# Effectiveness of vaccination against SARS-CoV-2 Omicron variant infection, symptomatic disease, and hospitalisation: a systematic review and meta-analysis

**DOI:** 10.1101/2022.06.23.22276809

**Authors:** Angela Meggiolaro, M. Sane Schepisi, Sara Farina, Carolina Castagna, Alessia Mammone, Andrea Siddu, Paola Stefanelli, Stefania Boccia, Giovanni Rezza

## Abstract

**Background:** The rapid rise of Sars-Cov2 B.1.1.529 variant (named Omicron) in the late November 2021 prompted the health authorities to estimate the potential impact on the existing countermeasures, including vaccines. This meta-analysis aims to assess the effectiveness of the current Sars-Cov2 vaccine regimens against laboratory-confirmed Omicron infection. A secondary endpoint aims to investigate the waning effectiveness of primary vaccination against symptomatic Omicron infection and related hospitalization.

**Methods:** The systematic review started on December 1, 2021 and was concluded on March 1, 2022. Random-effects (RE) frequentist meta-analyses are performed to estimate the primary vaccination course and the booster dose effectiveness against Omicron. Multiple meta-regressions are performed under mixed-effects model. This study is registered with PROSPERO, CRD42021240143.

**Findings:** In total, 15 out of 502 records are included in the quantitative synthesis. The meta-analysis on B.1.1.529 infection risk produces an OR=0·69 (95%CI: 0·57 to 0·83; τ2=0·225; I2=99·49%) after primary vaccination and an OR=0·30 (95%CI: 0·23 to 0·39; τ2=0·469; I2=99·33%) after one additional booster dose. According to the multiple meta-regression models, one booster dose significantly decreases by 69% the risk of symptomatic Omicron infection (OR=0·31; 95%CI: 0·23 to 0·40) and by 88% the risk of hospitalization (OR=0·12; 95%CI: 0·08 to 0·19) with respect to unvaccinated. Six months after primary vaccination, the average risk reduction declines to 22% (OR=0·78; 95%CI: 0·69 to 0·88) against symptomatic infection and to 55% against hospitalization (OR=0·45; 95%CI: 0·30 to 0·68).

**Interpretation:** Despite the high heterogeneity, this study confirms that primary vaccination does not provide sufficient protection against symptomatic Omicron infection. Although the effectiveness of the primary vaccination against hospitalization due to Omicron remains significantly above 50% after 3 months, it dramatically fades after 6 months. Therefore, the administration of one additional booster dose is recommended within 6 months and provides a 76% decrease in the odds of symptomatic Omicron after five months.

**Funding:** There was no funding source for this study.

**ARTICLE HIGHLIGHTS:**

- the primary vaccination decreases the risk of Omicron infection by 31%, while one additional booster dose decreases the risk by 70%
- the primary vaccination course reduces the risk of symptomatic Omicron infection by 24% and the risk of hospitalization by 50%
- one additional booster dose decreases by 69% the risk of symptomatic Omicron infection and the risk of hospitalization by 88%
- the effectiveness of the primary vaccination against hospitalization dramatically wanes after 3 months from vaccination, reaching a minimum of 45% in risk reduction after more than 6 months

**PANEL: research in context:** *Evidence before this study:* Omicron variant’s higher transmissibility combined with an increased risk of infection among individuals vaccinated with primary vaccination have prompted health authorities to introduce a booster vaccination. The systematic review including “vaccine effectiveness”, “Covid-19”, “SARS-CoV-2”, and “Omicron” search terms, is performed over three web engines and one early stage research platform (i.e., WHO COVID-19 DATABASE, PubMed, medRxiv + bioRxiv) Additionally, all relevant web sources reporting living data on vaccine effectiveness (i.e., https://view-hub.org/covid-19/ and https://covid-nma.com/), electronic databases and grey literature are considered. The last search update was on March 1, 2022. No country, language, study design restrictions are applied.

*Added value of this study:* Primary vaccination provides relatively low protection against the Omicron VOC, while one additional booster dose decreased substantially the risk of symptomatic Omicron infection and of hospitalization.

*Implications of all the available evidence:* The booster dose should be recommended after three months and no later than six months after the primary course vaccination, in order to avoid severe consequences, in particular among the elderly population.

## Introduction

On 26 November 2021, WHO designated the variant B.1.1.529 (named Omicron) as a variant of concern. (1)[WHO 1] The global epidemiology of SARS-CoV-2 has been characterized by the rapid spreading of the Omicron variant (B.1.1.529) ever since and Omicron has become the dominant variant circulating globally. (1)**[GISAID]** To date, Omicron encompasses several sub lineages, the most common ones are BA.1, BA.1.1 and BA.2.

The SARS-CoV-2 Omicron variant contains several important mutations on the spike protein, potentially leading to deleterious consequences. The increased transmissibility of Omicron is determined by a combination of: i) intrinsic biological properties that make the virus more infectious than previous lineages (e.g. ACE2 receptor binding efficiency or viral replication efficiency) (2)(3)**[Peacock, Abdelnabi]**; and ii) immune escape properties resulting in more breakthrough infections among vaccinated or more reinfections among recovered individuals.(4) (5)**[Viana, Yang]**

Regarding the clinical severity, a less severe onset, lower hospital admission rates and/or shorter length of hospital stay, as well as declining case fatality rates have been extensively documented by the scientific literature.(6) (7)(8)(9)**[Ferguson, Bager, Lewnard, Nyberg]**

COVID-19 vaccines licensed in the EU have proven highly effective in preventing SARS-CoV-2 infections (10) (11)(12)(13)(14)**[Hayawi, Harder, Liu, Kow, Pormohammad]**, however, several in vitro studies suggest a reduction in neutralizing titres against Omicron in individuals who have received vaccination with two or three doses and in those who have had prior SARS-CoV-2 infection.(15) (16)(17)**[Cele, Wilhelm, Scmidt]** Clinical studies have suggested that the levels of antibodies after BNT162b2, mRNA-1273 and Ad26.COV2.S vaccines could last for at least 6 months and decrease over time thereafter. (18)(19)(20)**[Barouch, Doria-Rose, Naaber]** Nonetheless, recent findings on cross-neutralizing immunity against Omicron among individuals that received a third dose of mRNA vaccine suggest that the current vaccine regimens may still overcome evasion of humoral immunity.(21)**[Garcia-Beltran]**

Omicron variant’s higher transmissibility combined with an increased risk of infection among vaccinated individuals, has prompted health authorities to consider the introduction of a booster dose. (22)**[Eroglu]** Therefore, estimating whether and how Sars-Cov2 primary vaccination effectiveness fades over time is essential to pinpoint the optimal timing for the booster dose.

The objective of this meta-analysis is twofold: first, to assess Sars-Cov2 vaccine effectiveness against infection, symptomatic disease, and hospitalization due to laboratory-confirmed SARS-CoV-2 Omicron variant. Second, to investigate the waning effectiveness of the primary course vaccination against Omicron over time.

## Methods

### Search strategy and selection criteria

This systematic review, with meta-analysis, is based on a web search updated weekly until March 1, 2022 **(Table S1, Supplementary material)**. The sources of information essentially consist of three web engines, including early stage research platforms (i.e., WHO COVID-19 DATABASE, PubMed, medRxiv + bioRxiv), all relevant web resources reporting living data on vaccine effectiveness (i.e., https://view-hub.org/covid-19/ and https://covid-nma.com/), electronic databases, and grey literature. Reviews and their references are examined for inclusion. No country, language, study design restrictions are applied.

All the relevant records are screened by title and abstract. Potentially relevant publications undergo full-text examination disagreements on eligibility are solved through discussion by all the authors. The full texts suitable for the quantitative synthesis are collected in an excel database for the data extraction. The items for data extraction are predefined and agreed upon by all authors. The systematic review and meta-analyses are performed in accordance with the Preferred Reporting Items for Systematic Reviews and Meta-Analyses (PRISMA) 2020 Statement guidelines. (23) **[Page]** (PRISMA checklist: Supplement 1) This study is registered with PROSPERO, CRD42021240143. (https://www.crd.york.ac.uk/PROSPERO/)

### Data extraction

Data extracted by at least three out of five independent investigators are collected in Excel tables. The information drawn up from each full text include:

1. General characteristics of the study: design, year of publication, country, mean age of the sample, follow-up, risk of bias;
2. Exposure: data are stratified according to Sars-Cov2 vaccination course, hence, two main groups are acknowledged, corresponding to primary vaccination and one additional booster dose recipients. Within each subgroup, the vaccination course is classified according to the vaccine type (ChAdOx1 nCoV-19, Ad26.COV2.S, BNT162b2, or mRNA-1273 vaccine). Heterologous primary schedules are included. All SARS-CoV-2 vaccine recipients are considered as *exposed*, while unvaccinated are considered as *unexposed*.
3. Outcome: cases are defined as being due to the Omicron variant, based on S target–negative results on PCR or whole-genome sequencing. Regardless the vaccine course undertaken, cases occurred within 14 days after the primary vaccination or within one week from the booster administration are not included. Omicron cases are classified by clinical severity into any Sars-Cov2 infection excluding hospitalization, symptomatic disease and hospitalization due to COVID-19 disease.
4. Risk of bias: ROBINS-I (risk of bias in non-randomized studies of interventions) is applied to assess risk of bias. The tool classifies the risk into “low”, “moderate” and “serious”. (24)**[Sterne]**

### Endpoints

The primary endpoint aims to assess the overall effectiveness of the current Sars-Cov2 vaccination regimens against Omicron. Study results are stratified by clinical severity and reported at the maximum follow-up and. The secondary endpoint attempts to measure the waning effectiveness of the primary vaccination at consecutive time intervals. In particular, VE is assessed in intervals of 3, 6, and more than 6 months after the last dose.

The point estimates of the effect size, as measured by Log odds ratios (Log ORs) and 95% confidence interval (95%CI), are computed through meta-analysis and converted to ORs by exponentiation. VE is quantified as the risk reduction of any infection event, expressed as a percentage, compared to the unvaccinated group.

### Statistical analysis

A random-effects (RE) model employing inverse variance method (IV) is fitted to the data. The amount of heterogeneity (i.e., τ^2^), is estimated using the restricted maximum-likelihood estimator. (25)**[Viechtbauer]** In addition to the estimate of τ^2^, the QQ-test for heterogeneity (26)**[Cochran]** and the I^2^ statistic (27)**[Higgins]** are reported. Studentized residuals and Cook’s distances are used to examine whether studies may be outliers and/or influential in the context of the model. The normality assumption is evaluated via QQ normal plot. (28)**[Viechtbauer 2]**

The publication bias is evaluated through a funnel plot and tested via regression test (weighted regression with multiplicative dispersion). The rank correlation test (29,30)**[Begg]** and the regression test (30)**[Sterne 2]**, using the standard error of the observed outcomes as predictor, are used to check for funnel plot asymmetry. Regarding the primary vaccination waning effectiveness, the subgroup meta-analyses include the stratification by time intervals since the last dose uptake for symptomatic Covid-19 risk and hospitalization risk due to Sars-Cov2 infection. Studies providing vaccine effectiveness estimates at discrete time intervals after the primary vaccination course, which met the predefined screening criteria, underwent further meta-analysis and meta-regression. In order to test for subgroup differences, both a mixed effect meta-regression model assuming a common τ^2^ value within the subgroups and a three level meta-regression model, allowing for different τ^2^ values across subgroups, are fitted.

Finally, in order to examine whether one or multiple moderator variables are able to account for the heterogeneity (or part of it), multiple meta-regressions are performed under the mixed-effects model for both continuous and nominal study level covariates.(25)**[Viechtbauer 2005]** The analysis is carried out using R (version 4.0.5).

## Results

The web search provided 502 unduplicated records. **(Figure 1)** In total, 15 studies and 55 observations are included in the quantitative synthesis concerning the overall Sars-Cov2 vaccine effectiveness against Omicron VOC. All of them have a test-negative case-control design except one cohort study. (31)**[Spensley] (Table S2, Supplementary material)**. The majority of the studies are carried out in US and UK (59%), the sample age is on average 45 years, while the induction period for immunization appears slightly shorter for studies analysing the effectiveness of the booster dose compared to those investigating the primary course vaccination (on average 11 and 16 days since administration, respectively). The 75% of the observations concern the mRNA vaccine effectiveness, while the 13% involve heterologous vaccine regimens. The booster dose is administered on average 6 months after the primary course **(Table S1, Supplementary material 2)**. The majority of the selected studies involves the general population, while **Gray** et al. report results on HCWs (32)**[Gray]** and **Spensley et al**. on patients affected by end stage kidney disease receiving in hospital haemodialysis. (31)**[Spensley]** All the studies examine the VE of mRNA BNT162b2 vaccine except Tseng et al. and Gray et al. which investigate mRNA-1273 and aAd26.COV.2 effectiveness, respectively. (33)(32)**[Tseng] [Gray]**

**Figure 1.**
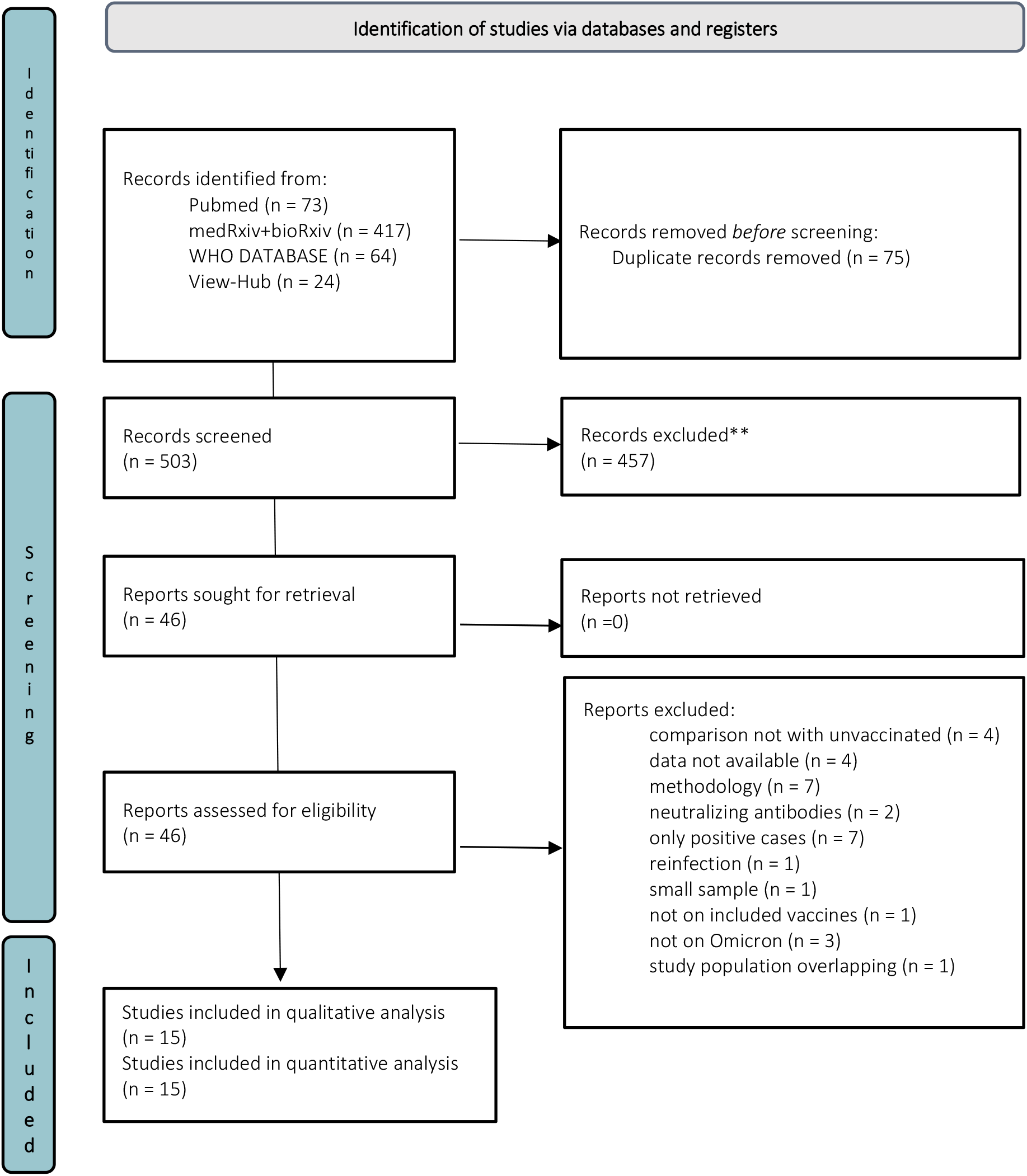
Prisma flow diagram.

Five studies report data on the waning effectiveness of the primary vaccination against symptomatic Omicron infection. (34)(35,36)(36)(37)(38)**[Accorsi, Andeweg, Chemaitelly, Powell, Sheikh]** A considerable effectiveness rebound after mRNA booster dose is shown by 4 studies.(34)(35)(36)(38)**[Accorsi, Andeweg, Chemaitelli, Sheikh]** VE against hospitalisation caused by Omicron is analysed by seven studies. (32)(33)(36)(39)(40)(41) (42)**[Gray, Tseng, Chemaitelly, Collie, Ferdinands, Gray, Tartof, Thompson]**

### Risk of Omicron infection after primary course vaccination

A total of 14 studies and k=27 observations are included in this meta-analysis. The median follow-up period is 213 days (70-365). The observed log odds ratios range from −1·275 to 0·467, with the majority of estimates being negative (67%). The estimated average log odds ratio based on the RE model is 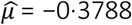 = −0·3788 (95% CI: −0·568 to −0·190). The values are transformed into the odds ratio scale through exponentiation, such that OR= 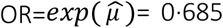 = 0·685 (95%CI: 0·567 to 0·827). The average outcome differs significantly from zero (z=−3·931, p<0·0001)· Hence, the result suggests that the risk of Sars-Cov2 infection in vaccinated individuals is on average 31·5% lower than the infection risk in unvaccinated. The forest plot is exhibited in. According to the Q-test, the true outcomes appear to be heterogeneous (Q (26) = 1962·9, p<0·0001; τ^2^=0·225; I^2^=99·49%). Neither the rank correlation nor the Egger’s regression test indicate any funnel plot asymmetry (p=0·901 and p=0·409, respectively). The analysis of heterogeneity is displayed in **Figures S1-S5 (Supplementary material)**.

The subgroup analysis includes five subgroups, three of them display significant results (p<0·05). Regarding the vaccines used for the primary vaccination, only messenger RNA (mRNA) vaccine exhibits a significant OR=0·62 (95%CI: 0·51 to 0·76). **(Figure S11, Supplementary material)**. The stratified meta-analysis assessing the primary vaccination effectiveness against Omicron VOC by severity of symptoms includes three subgroups and the test for subgroup differences is significant (Q_M_(df = 2) = 23·30, p < 0·0001) **(Figure 2)**. According to the three level meta-analysis, the 35·6% of the total variance is distributed within effect sizes (second level), whilst the 64·1% is distributed between groups (third level). The multiple meta-regression embeds four moderators: risk of bias, mean age of the samples (variable centred on the overall mean value of 45 years), vaccine employed in the primary course vaccination (viral vector vaccine or ‘VV’, mRNA and heterologous vaccination with both VV and mRNA or ‘VV/mRNA’). Albeit reduced, the residual heterogeneity remains significant (QE (df=19) =448·6, p<0·0001); τ^2^=0·0701; I^2^=97·25%). The (0.2254 -0.0701)/ 0.2254 = 68.9% of the total amount of heterogeneity can be explained by including four moderators in the meta-regression, suggesting further unobserved effect not captured by the model. On average, the risk of symptomatic Covid-19 appears 24% lower for the vaccinated group compared to the unvaccinated (OR=0·76; 95%CI: 0·58 to 0·99), while the risk of hospitalization is 50% lower for the vaccinated group (OR=0·50; 95%CI: 0·34 to 0·72). The OR estimate for any positive rt-PCR is not significant. **(Figure S13 and Table S4, Supplementary material)**.

**Figure 2.**
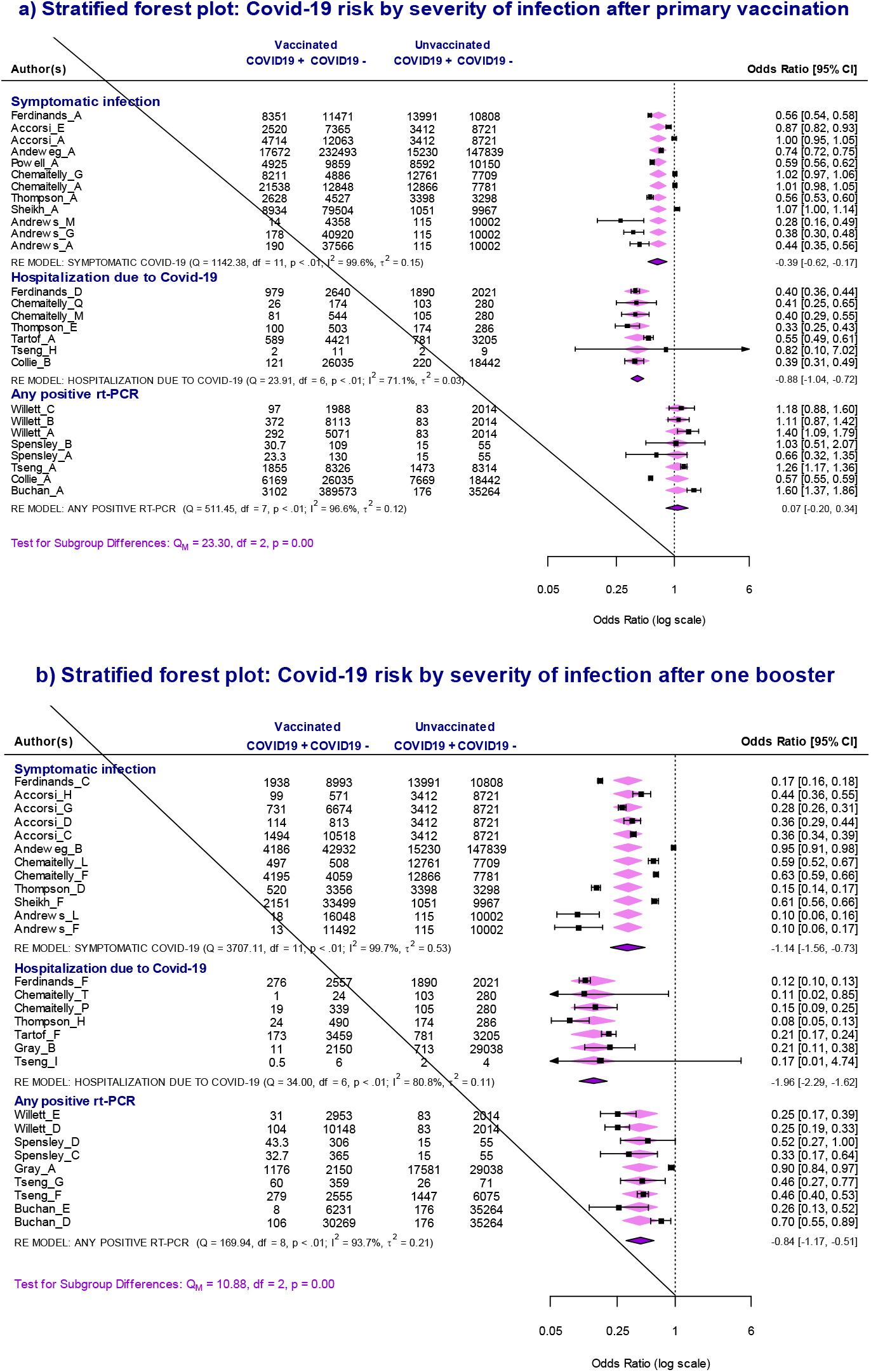
Stratified forest plots and subgroup meta-analyses. Random effect model, IV method. (a) Effectiveness of primary course vaccination, by severity of symptoms. The risk of symptomatic Covid-19 is assessed by 12 observations, the risk of hospitalization by seven and the risk of any positive rt-PCR by eight. The test for subgroup difference is significant (QM (df =2) =23·3, p<0·0001). According to the subgroup analysis, the risk of any positive rt-PCR appears 7% higher among the vaccinated group with respect to the unvaccinated, however, the result is not significant (OR= 1·07; 95%CI: 0·82 to 1·40). The risk reduction for symptomatic Covid-19 is 32% lower among the vaccinated group compared to the unvaccinated (OR=0·68; 95%CI: 0·54 to 0·85). Regarding hospitalization due to Omicron infection, the risk appears 58% lower for the vaccinated group compared to unvaccinated (OR= 0·42; 95%CI: 0·35 to 0·49). (b) Effectiveness of one booster dose against Omicron VOC, by severity of symptoms. The effectiveness of one booster dose is estimated by 12 observations for the symptomatic Covid-19, by seven for hospitalization risk and by nine for any positive rt-PCR. The test for subgroup difference is significant (QM (df =2) =10·88, p<0·0001). The risk of positive rt-PCR appears 57% lower among the booster group with respect to the unvaccinated group (OR=0·43; 95%CI: 0·31 to 0·60). The risk reduction in favour of the booster group is 68% for symptomatic Covid-19 (OR=0·32; 95%CI: 0·21 to 0·48) and 86% for hospitalization (OR=0·14; 95%CI: 0·10 to 0·20).

**Figure 3.**
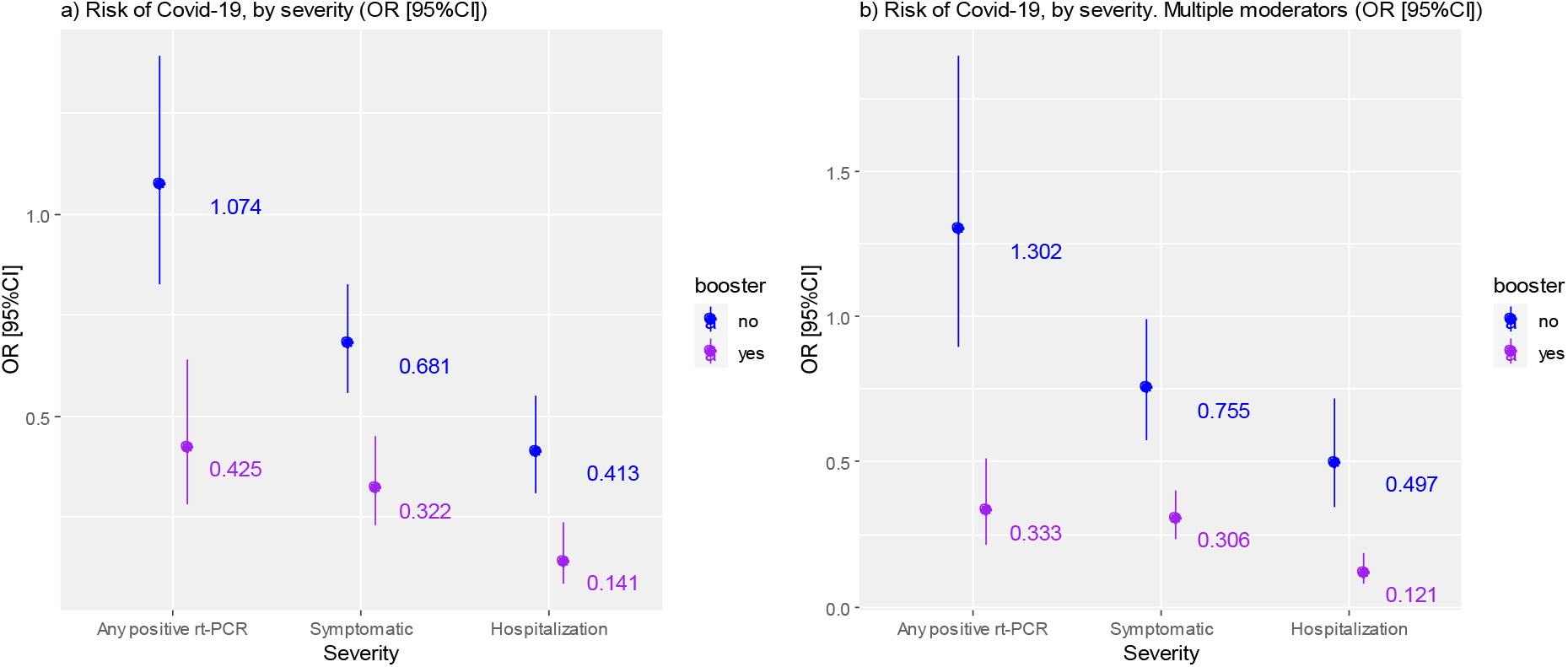
Meta-regression model estimates, risk of Omicron infection by severity of symptoms: OR [95%CI] estimates from meta-regression with one moderator (a) and multiple moderator (b). In the restricted meta-regression (one moderator), the risk of any positive rt-PCR appears 7% higher among the primary vaccination group with respect to unvaccinated (OR= 1·07; 95%CI: 0·83 to 1·39); whilst in the multiple meta-regression, the risk of any positive rt-PCR appears nearly 30% higher for primary vaccination. However, the results are not significant (OR= 1·302; 95%CI: 0·894 to 1·898).

### Risk of Omicron infection after one booster dose

A total of k=28 observations and 13 studies are included in this meta-analysis. The median follow-up is 62 days (14-150). All the studies investigate the effectiveness of mRNA booster dose except ‘Gray’, which demonstrates the efficacy of a two-dose regimen of Ad26.COV.2 vaccine.(32) **[Gray]** The observed log odds ratios range from −2·5194 to −0·0550, with the 100% of estimates being negative. The average log odds ratio based on the RE model is 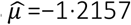 =−1·2157 (95%CI: −1·4854 to −0·9460). Therefore, the outcome differs significantly from zero (z=−8·8351, p<0·0001). The exponentiation yields an average OR=0·296 (95%CI: 0·226 to 0·388), hence, vaccinated with one booster dose have a 70·4% risk reduction of Omicron infection compared to unvaccinated. According to the Q-test, the true outcomes appear to be heterogeneous (Q (27) =4624·51, p<0·0001; τ^2^=0·4686; I^2^=99·33%). The influential analysis does not detect overly influential outliers. **(Figure S6-S10, Supplementary material)**. There is no indication of publication bias because neither the rank correlation nor the regression test indicates any funnel plot asymmetry (p=0·7992 and p=0·0735, respectively). The subgroup analysis includes six subgroups, four of them display significant results (p<0·05). Notably, the risk reduction for the booster group seems 69% lower in studies reporting results at 3 months of follow-up (OR= 0·31; 95%CI: 0·23 to 0·42) and 76% in studies reporting 5 months follow-up (OR= 0·24; 95%CI: 0·12 to 0·428) at most. However, the test for interaction is not significant **(Figure S12, Supplementary material)**.

The stratified meta-analysis on one booster effectiveness against Omicron VOC by clinical severity includes three subgroups **(Figure 2b)**. The test for subgroup differences is significant (QM (df=2) = 10·88, p=0·004). According to the multilevel meta-analysis approach, the 57·3% of the total variance is distributed within effect sizes at second level, whilst the 42·2% is distributed between groups (level 3).

The multiple meta-regression model includes four moderators: risk of bias, mean age of the samples (centred on the overall mean value of 44·43 years), regimen of the primary course vaccination (‘VV’, mRNA, ‘VV/mRNA’). As expected, the residual heterogeneity slightly decreases but remains significant (QE (df = 20) =306·9, p<0·0001; τ^2^=0·1037; I^2^=94·54%) suggesting further unobserved effect not captured by the predictors in the model. Overall, the multiple meta-regression can explain the 77·9% of the total amount of heterogeneity. On average, the risk of symptomatic Covid-19 appears, 69% lower for the booster group compared to the unvaccinated (OR=0·31; 95%CI: 0·23 to 0·40), whilst the risk of hospitalization is on average 88% lower (OR=0·12; 95%CI: 0·08 to 0·19) **(Figure S14 and Table S4, Supplementary material)**.

### Waning effectiveness of Sars-Cov2 primary vaccination against Omicron VOC

Overall, eight studies assess the effectiveness of the primary vaccination against Sars-Cov2 at consecutive time intervals. The time intervals correspond to three months, three to six months, six months and longer than six months since the last dose administration. Therefore, the stratified meta-analyses on Sars-Cov2 vaccine waning effectiveness against Omicron include four subgroups. **(Figure 4)** The risk of developing a symptomatic Covid-19 is investigated by seven studies and the risk of hospitalization is investigated by four studies (Table S5, Supplementary material) Only in 33% of cases, Omicron rt-PCR positivity is tested routinely, therefore, it is not possible to consistently estimate the vaccine effectiveness in preventing Sars-Cov2 infection, as well as the vaccine’s capability of limiting the virus spreading.

**Figure 4.**
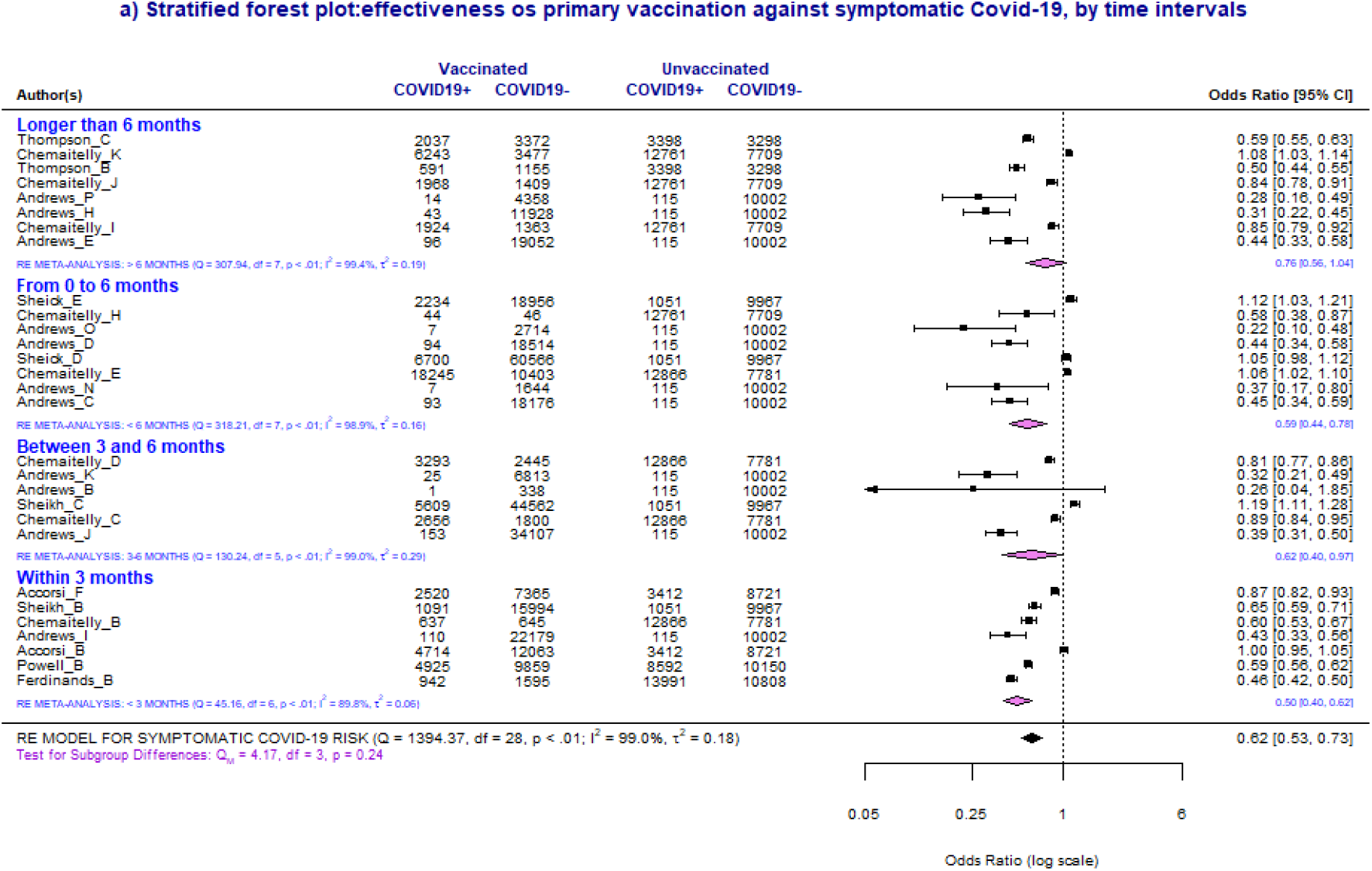
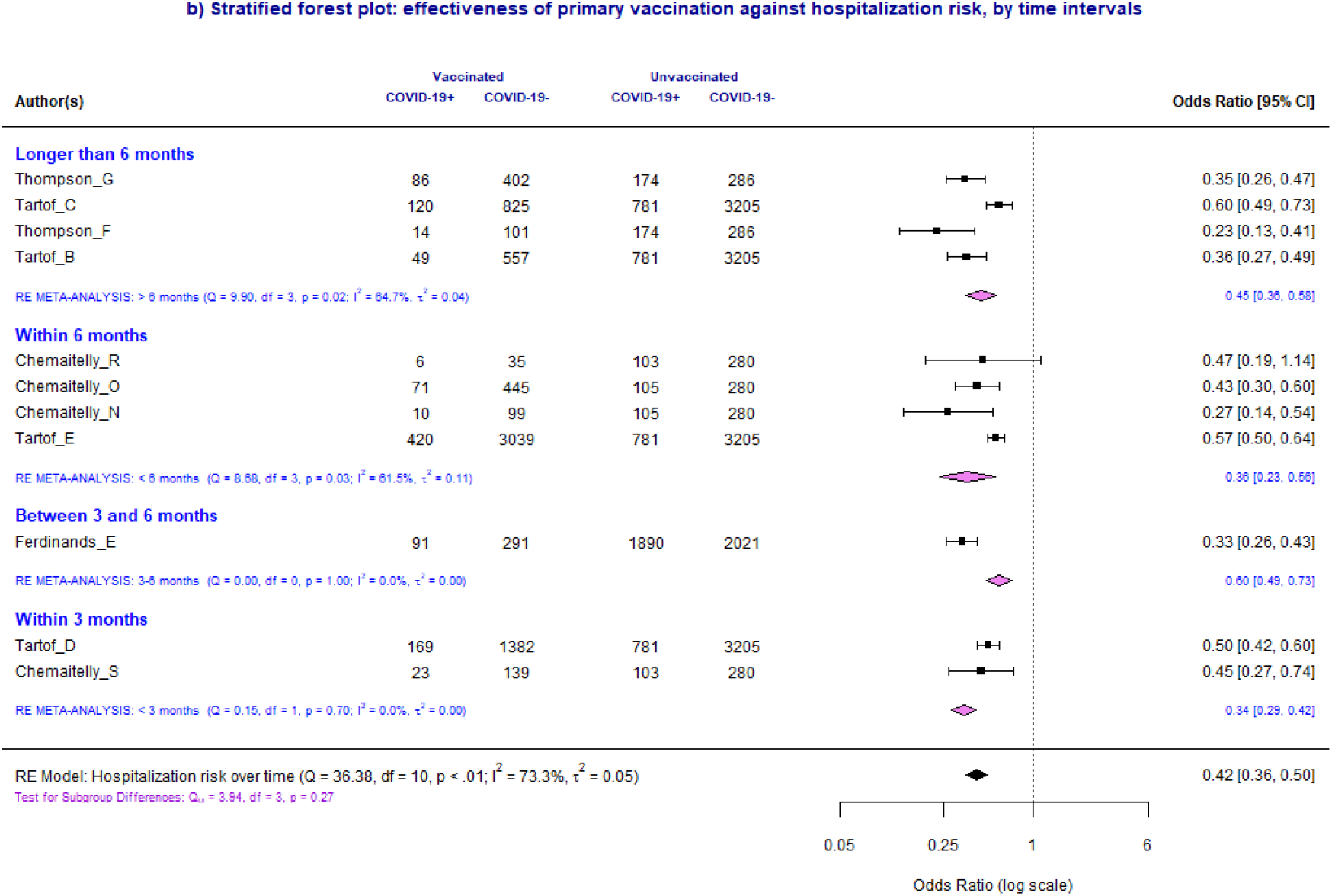
Stratified Forest Plots. The forest plots include four subgroups representing four discrete time intervals. The results of the individual studies are grouped together according to the corresponding subgroup. Below each subgroup, a summary polygon shows the results of a RE meta-analysis. The pooled effect sizes are expressed as Log Odds Ratios. The summary polygon at the bottom of the plot shows the results from the overall RE model (IV method). (a) Stratified forest plot, symptomatic Omicron infection risk, by time intervals. According to the subgroup analysis, the risk reduction appears to be 50% among vaccinated compared to unvaccinated until 3 months (OR=0·50; 95%CI: 0·40 to 0·62). The risk reduction decreases to nearly 41% with respect to unvaccinated within 6 months (OR=0·59; 95%CI: 0·44-0·78), and to 24% thereafter (OR=0·76; 95%CI: 0·56 to 1·03). (b) Stratified forest plot, hospitalization due to Omicron infection risk, by time intervals. The risk reduction appears 65% lower among vaccinated compared to unvaccinated within 3 months (OR=0·35; 95%CI: 0·29 to 0·42); whereas the overall risk reduction is on average 64% compared to unvaccinated within 6 months (OR=0·36; 95%CI: 0·23 to 0·56) and 54% thereafter (OR=0·46; 95%CI=0·36 to 0·58). Only one study assesses the risk of hospitalization between 3 and 6 months (OR=0·60; 95%CI=0·49 to 0·73) (41)**[Tartof]**

Concerning the risk of symptomatic Omicron infection after vaccination with primary course, a total of k=29 observations are included in the meta-analysis; all estimates are based on the RE model. The overall Log odds ratio based on the RE model is 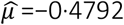 =−0·4792 (95% CI: −0·6418 to −0·3165), equivalent to OR=0·62 (95%CI=0·53-0·73) after exponentiation. The outcomes appear heterogeneous (Q (28) =1394·37, p<0·0001; τ^2^=0·1773; I^2^=99·01%) and the regression test indicates funnel plot asymmetry (p<0·0001), however, the rank correlation test is not significant (p=0·3051) **(Figure S17a, Supplementary material)**. The test for subgroup differences is not statistically significant (Q_M_ (df =3) =4·169, p=0·2438) **(Figure 4a)**. According to the multilevel meta-analysis, the 93·1% of the total variance is distributed at second level (σ^2^=0·168) while the 5·9% is distributed at third level (between groups).

The multiple meta-regression model includes four moderators: time-lapse since the last dose, risk of bias, age of the study (variable centred on the mean value of 41.4), vaccine technology (VV, mRNA, heterologous vaccination or VV/mRNA). The residual heterogeneity notably decreases but remains significant (QE (df=20) = 53·6, p<0·0001; τ^2^=0·0042; I^2^=65·10%). The average risk reduction is 46% for vaccinated with respect to unvaccinated (OR=0·54; 95%CI: 0·48 to 0·61) within 3 months, 22% within 6 months (OR=0·78; 95%CI: 0·69 to 0·88) and 16% between 3 and 6 months (OR=0·84; 95%CI: 0·74 to 0·96). Moreover, the OR decreases on average by 2% in studies where the mean age is one more unit away from the overall mean of 41·4 years (OR=0·98; 95%CI: 0·98 to 0·99). The heterologous vaccination (VV/mRNA) provides a positive coefficient and an 18% higher risk of symptomatic Omicron infection with respect to mRNA vaccine regimens (OR=1·18; 95%CI: 1·08 to 1·29) **(Figure S20 and Table S6, Supplementary material)**.

The meta-analysis on Sars-Cov2 primary vaccination effectiveness against hospitalization embeds a total of 4 studies and 11 observations **(Figure 4b)**. The average log odds ratio based on the RE model is 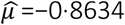 =−0·8634 (95%CI: −1·0348 to −0·6920), which corresponds to OR=0·42 (95%CI: 0·36 to 0·50) by exponentiation. According to the Q-test, the true outcomes appear heterogeneous (Q (10)=36·38, p<0·0001; τ^2^=0·051; I^2^=73·3%). The regression test indicates funnel plot asymmetry (p=0·0272); however it is not confirmed by the rank correlation test (p=0·542) **(Figure S17, Supplementary material)**. The test for subgroup differences suggests that there is not a statistically significant subgroup effect (Q_M_ (df =3) =3·9437, p=0·268). The three level meta-analysis approach shows that the 73·3% of the total variance is distributed at second level (σ^2^=0·051) while σ^2^=0·00 at third level.

In the multiple meta-regression model, only three predictors are designated as moderators because ‘vaccine regimen’ contains only observations on mRNA vaccines. The estimated amount of residual heterogeneity is τ^2^=0·00 and the test for residual heterogeneity is no longer significant (QE (df =4) =1·83, p=0·766). All moderators exhibit significant coefficients except ‘risk of bias’. The adjusted average effect corresponds to an OR=0·28 (95%CI= 0·21 to 0·38) within 3 months and average risk reduction of 72% for vaccinated in comparison to unvaccinated. The average OR raises to 0·38 (95%CI: 0·25 to 0·59) within 6 months and to 0·45 (95%CI: 0·30 to 0·68) after more than 6 months. The variable ‘age’ (centered on the mean of 48·3 years) generates a significant coefficient indicating that the risk of hospitalization increases on average by 2·6% by increasing of one unit the study mean age (OR=1·026; 95%CI: 1·003 to 1·049) **(Figure S21 and Table S6, Supplementary material)**. The predicted ORs for symptomatic infection and hospitalization risks are plotted in **Figure 5**.

**Figure 5.**
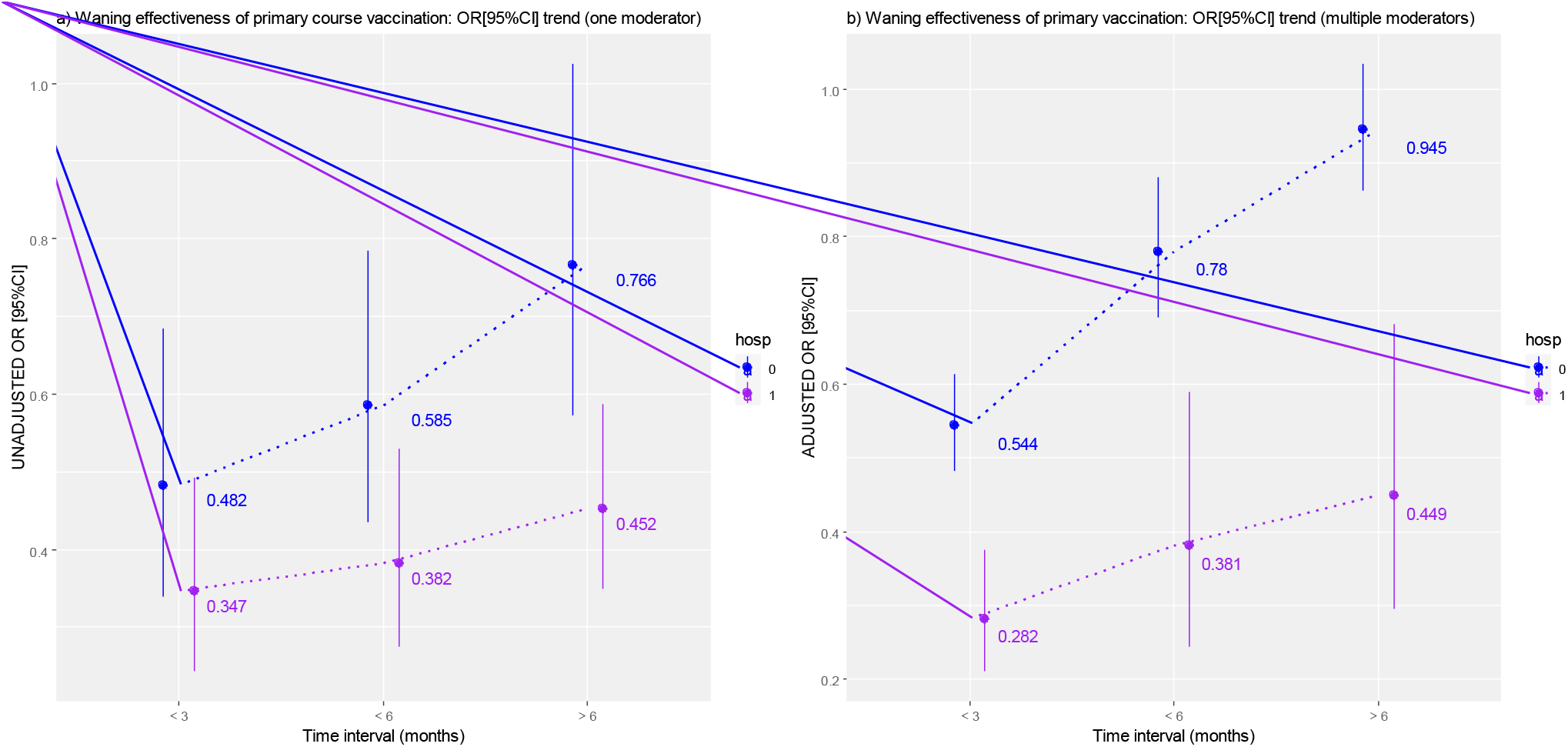
Plots displaying the trend of symptomatic Omicron infection and related hospitalization risk. Y-axes: ORs [95%CI] estimates from meta-regression with one moderator (a) and meta-regression with multiple moderators (b). The risk of symptomatic Covid-19 infections is depicted in blue while the risk of hospitalization in purple. X-axes: time intervals at 3 months, 6 months and over 6 months from last dose administration. Time interval running from 3 to 6 months is suppressed because only one study estimate is available for hospitalization risk.

## Discussion

The evidence achieved through the quantitative synthesis suggests that a primary vaccination course is not sufficiently protective against Omicron. In fact, the probabilities of symptomatic infection and related hospitalization are nearly 50% for vaccinated with respect to unvaccinated, basing on a maximum follow up of one year. One additional booster dose decreases by 69% the risk of symptomatic Omicron infection (OR=0·31; 95%CI: 0·23 to 0·40) and by 88% the risk of hospitalization (OR=0·12; 95%CI: 0·08 to 0·19) with respect to unvaccinated at a maximum follow-up of five months. Albeit not significant, the subgroup analysis does not suggest a waning effectiveness of the booster dose after five months, however, the evidence on long term effectiveness is still limited.

The risk of any positive rt-PCR appears higher among the primary vaccination group with respect to the unvaccinated (OR= 1·302; 95%CI: 0·89 to 1·90); however, the results are not significant.

Age does not appear as a significant predictor, notwithstanding the negative association with the overall risk of infection after the primary vaccination and after the booster. Conversely, age is negatively associated with the risk of symptomatic infection and positively associated with the risk of hospitalization after primary vaccination. Some unobserved effect of uncontrolled confounding must be acknowledged in interpreting this association. For instance, the different extent to which the joint effect of the mitigation measures uplift has affected the younger and the elderly population. However, despite the generalizability, these results do not allow to infer any clear conclusion.

There is no clear advantage between homologous and heterologous vaccination, particularly on boosting, probably because the majority of the appraisals have been conducted on mRNA vaccination and data on heterologous vaccination are quite sparse.

As the administration of booster doses, whether homologous or heterologous, should take into consideration the waning protection of the primary course and the optimal interval for an efficient immune response, the implications of our findings extend to health care and public health policy.

Our results on the waning trends align with the estimates provided by the clinical trials. (43)(44)(45)**[Polack, Baden, Sadoff]** According to our estimates, the effectiveness of primary vaccination against Omicron reaches a peak within three months determining a risk reduction of roughly 72% with respect to unvaccinated. The protection is maintained at six months, with risk reduction of nearly 62%, and dramatically decline thereafter (55% less probabilities for vaccinated compared to unvaccinated). Overall, the effectiveness against hospitalization diminishes by approximately 10-15% every three months, and the point estimates show wide confidence intervals. (46)**[Lin]**

The ramping-up trend for symptomatic Omicron infection risk appears steeper than the trend for hospitalization risk; in other words, the protection against the symptomatic Covid-19 declines faster. The risk reduction of symptomatic Omicron infection after a primary vaccination declines sharply to 22% in six months. Our study provides the best available data synthesis on vaccine effectiveness against Omicron; however, several limitations must be acknowledged. First, only in 33% of cases, Omicron rt-PCR positivity is tested routinely, therefore, it is not possible to draw conclusion on vaccine effectiveness in preventing Omicron infection. Second, by examining periods during which Omicron and Delta coexistence was very likely, early studies generate a distortion of the VE effectiveness estimate.

In part, the high heterogeneity surrounding the meta-analysis estimates stems from the observational design of the included studies. Unless a randomization process, the meta-regression cannot capture the unobserved effect of confounders such as the level of community transmission, the implementation of public health prevention measures, and the surge of new variants. For instance, regarding the Omicron variant definition, the studies on BA.1 do not distinguish between the different sub-lineages, although the majority of them are conducted during the BA.1 surge. Differences between BA.2 and BA.1 in evading immunity remain undefined. In conclusion, despite the high heterogeneity, only in part explained by the meta-regressions, this study confirms that primary vaccination does not provide sufficient protection against symptomatic Omicron infection, because the overall effectiveness estimate never reaches a minimum requirement of 50% in risk reduction. One additional booster dose decreases substantially the risk of symptomatic Omicron infection and of hospitalization. The booster dose administration should be recommended after three months and no later than six months following the primary vaccination course.

## Data Availability

All data produced in the present study are available upon reasonable request to the authors.

## Contributors

Conceptualization, G.R., A.M. and M.S.S.; methodology, M.S.S. and A.M.; formal analysis, A.M.; data curation, A.M., M.S.S., A.M., C.C., S.F., A.S.; writing—original draft preparation, A.M. and M.S.S.; writing—review and editing, A.M., C.C., S.B., A.S, P.S. and M.S.S.; supervision, S.B. and G.R. All authors have read and agreed to the published version of the manuscript.

## Data sharing

Data supporting the reported results are available on request to the Authors.

## References

1. GISAID - hCov19 Variants [Internet]. [cited 2022 Jun 7]. Available from: https://www.gisaid.org/hcov19-variants/

2. Peacock 1# TP, Brown JC, Zhou J, Thakur N, Newman J, Kugathasan R, et al. The SARS-CoV-2 variant, Omicron, shows rapid replication in human primary nasal epithelial cultures and efficiently uses the endosomal route of entry. [cited 2022 Jun 7]; Available from: https://doi.org/10.1101/2021.12.31.474653

3. Abdelnabi R, Foo CS, Zhang X, Lemmens V, Maes P, Slechten B, et al. The omicron (B.1.1.529) SARS-CoV-2 variant of concern does not readily infect Syrian hamsters. bioRxiv [Internet]. 2021 Dec 26 [cited 2022 Jun 7];2021.12.24.474086. Available from: https://www.biorxiv.org/content/10.1101/2021.12.24.474086v1

4. Viana R, Moyo S, Amoako DG, Tegally H, Scheepers C, Althaus CL, et al. Rapid epidemic expansion of the SARS-CoV-2 Omicron variant in southern Africa. Nature [Internet]. 2022 Mar 24 [cited 2022 Jun 7];603(7902):679–86. Available from: https://pubmed.ncbi.nlm.nih.gov/35042229/

5. Yang W, Shaman J. SARS-CoV-2 transmission dynamics in South Africa and epidemiological characteristics of the Omicron variant. medRxiv [Internet]. 2021 Dec 21 [cited 2022 Jun 7]; Available from: http://www.ncbi.nlm.nih.gov/pubmed/34981071

6. [PDF] Report 50: Hospitalisation risk for Omicron cases in England | Semantic Scholar [Internet]. [cited 2022 Jun 7]. Available from: https://www.semanticscholar.org/paper/Report-50%3A-Hospitalisation-risk-for-Omicron-cases-Ferguson-Ghani/8b696e1a05092a11b39b92b4adea7bc05e7032df

7. Bager P, Wohlfahrt J, Bhatt S, Edslev SM, Sieber RN, Ingham AC, et al. Reduced Risk of Hospitalisation Associated With Infection With SARS-CoV-2 Omicron Relative to Delta: A Danish Cohort Study. SSRN Electronic Journal [Internet]. 2022 Jan 20 [cited 2022 Jun 7]; Available from: https://papers.ssrn.com/abstract=4008930

8. Lewnard JA, Hong VX, Patel MM, Kahn R, Lipsitch M, Tartof SY. Clinical outcomes among patients infected with Omicron (B.1.1.529) SARS-CoV-2 variant in southern California. [cited 2022 Jun 7]; Available from: https://doi.org/10.1101/2022.01.11.22269045

9. Nyberg T, Ferguson NM, Nash SG, Webster HH, Flaxman S, Andrews N, et al. Comparative analysis of the risks of hospitalisation and death associated with SARS-CoV-2 omicron (B.1.1.529) and delta (B.1.617.2) variants in England: a cohort study. The Lancet [Internet]. 2022 Apr 2 [cited 2022 Jun 7];399(10332):1303–12. Available from: http://www.thelancet.com/article/S0140673622004627/fulltext

10. Hayawi K, Shahriar S, Serhani MA, Alashwal H, Masud MM. Vaccine versus variants (3Vs): Are the COVID-19 vaccines effective against the variants? A systematic review. Vaccines (Basel). 2021 Nov 1;9(11).

11. Harder T, Külper-Schiek W, Reda S, Treskova-Schwarzbach M, Koch J, Vygen-Bonnet S, et al. Effectiveness of COVID-19 vaccines against SARS-CoV-2 infection with the Delta (B.1.617.2) variant: second interim results of a living systematic review and meta-analysis, 1 January to 25 August 2021. Euro surveillance : bulletin Europeen sur les maladies transmissibles = European communicable disease bulletin [Internet]. 2021 Oct 14 [cited 2022 Jun 7];26(41). Available from: https://pubmed.ncbi.nlm.nih.gov/34651577/

12. Liu Q, Qin C, Liu M, Liu J. Effectiveness and safety of SARS-CoV-2 vaccine in real-world studies: a systematic review and meta-analysis. Infectious Diseases of Poverty [Internet]. 2021 Dec 1 [cited 2022 Jun 7];10(1). Available from: /pmc/articles/PMC8590867/

13. Kow CS, Ramachandram DS, Hasan SS. The effectiveness of mRNA-1273 vaccine against COVID-19 caused by Delta variant: A systematic review and meta-analysis. J Med Virol [Internet]. 2022 May 1 [cited 2022 Jun 7];94(5):2269–74. Available from: https://pubmed.ncbi.nlm.nih.gov/34978339/

14. Pormohammad A, Zarei M, Ghorbani S, Mohammadi M, Neshin SAS, Khatami A, et al. Effectiveness of COVID-19 Vaccines against Delta (B.1.617.2) Variant: A Systematic Review and Meta-Analysis of Clinical Studies. Vaccines (Basel) [Internet]. 2021 Jan 1 [cited 2022 Jun 7];10(1). Available from: https://pubmed.ncbi.nlm.nih.gov/35062684/

15. Cele S, Jackson L, Khoury DS, Khan K, Moyo-Gwete T, Tegally H, et al. SARS-CoV-2 Omicron has extensive but incomplete escape of Pfizer BNT162b2 elicited neutralization and requires ACE2 for infection. medRxiv [Internet]. 2021 Dec 17 [cited 2022 Jun 7]; Available from: http://www.ncbi.nlm.nih.gov/pubmed/34909788

16. Wilhelm A, Widera M, Grikscheit K, Toptan T, Schenk B, Pallas C, et al. Reduced Neutralization of SARS-CoV-2 Omicron Variant by Vaccine Sera and monoclonal antibodies. medRxiv [Internet]. 2021 Dec 8 [cited 2022 Jun 7];2021.12.07.21267432. Available from: https://www.medrxiv.org/content/10.1101/2021.12.07.21267432v1

17. Schmidt F, Muecksch F, Weisblum Y, Silva J da, Bednarski E, Cho A, et al. Plasma neutralization properties of the SARS-CoV-2 Omicron variant. medRxiv [Internet]. 2021 Dec 13 [cited 2022 Jun 7];2021.12.12.21267646. Available from: https://www.medrxiv.org/content/10.1101/2021.12.12.21267646v1

18. Barouch DH, Stephenson KE, Sadoff J, Yu J, Chang A, Gebre M, et al. Durable Humoral and Cellular Immune Responses Following Ad26.COV2.S Vaccination for COVID-19. medRxiv [Internet]. 2021 Jul 7 [cited 2022 Jun 8];2021.07.05.21259918. Available from: https://www.medrxiv.org/content/10.1101/2021.07.05.21259918v1

19. Doria-Rose N, Suthar MS, Makowski M, O’Connell S, McDermott AB, Flach B, et al. Antibody Persistence through 6 Months after the Second Dose of mRNA-1273 Vaccine for Covid-19. N Engl J Med [Internet]. 2021 Jun 10 [cited 2022 Jun 7];384(23):2259–61. Available from: https://pubmed.ncbi.nlm.nih.gov/33822494/

20. Naaber P, Tserel L, Kangro K, Sepp E, Jürjenson V, Adamson A, et al. Dynamics of antibody response to BNT162b2 vaccine after six months: a longitudinal prospective study. The Lancet regional health Europe [Internet]. 2021 Nov 1 [cited 2022 Jun 7];10. Available from: https://pubmed.ncbi.nlm.nih.gov/34514454/

21. Garcia-Beltran WF, st. Denis KJ, Hoelzemer A, Lam EC, Nitido AD, Sheehan ML, et al. mRNA-based COVID-19 vaccine boosters induce neutralizing immunity against SARS-CoV-2 Omicron variant. Cell [Internet]. 2022 Feb 3 [cited 2022 Jun 7];185(3):457–466.e4. Available from: https://pubmed.ncbi.nlm.nih.gov/34995482/

22. Eroglu B, Nuwarda RF, Ramzan I, Kayser V. A Narrative Review of COVID-19 Vaccines. Vaccines (Basel) [Internet]. 2021 Jan 1 [cited 2022 Jun 7];10(1). Available from: https://pubmed.ncbi.nlm.nih.gov/35062723/

23. Page MJ, McKenzie JE, Bossuyt PM, Boutron I, Hoffmann TC, Mulrow CD, et al. The PRISMA 2020 statement: an updated guideline for reporting systematic reviews. BMJ [Internet]. 2021 Mar 29 [cited 2022 Jun 7];372. Available from: https://pubmed.ncbi.nlm.nih.gov/33782057/

24. Sterne JA, Hernán MA, Reeves BC, Savović J, Berkman ND, Viswanathan M, et al. ROBINS-I: a tool for assessing risk of bias in non-randomised studies of interventions. BMJ [Internet]. 2016 Oct 12 [cited 2022 Jun 7];355. Available from: https://www.bmj.com/content/355/bmj.i4919

25. Viechtbauer W. Bias and efficiency of meta-analytic variance estimators in the random-effects model. Journal of Educational and Behavioral Statistics. 2005;30(3):261–93.

26. Cochran WG. The Combination of Estimates from Different Experiments. Biometrics [Internet]. 1954 Mar [cited 2022 Jun 7];10(1):101. Available from: /record/1955-00087-001

27. Higgins JPT, Thompson SG. Controlling the risk of spurious findings from meta-regression. Stat Med [Internet]. 2004 Jun 15 [cited 2022 Jun 7];23(11):1663–82. Available from: https://pubmed.ncbi.nlm.nih.gov/15160401/

28. Viechtbauer W, Cheung MWL. Outlier and influence diagnostics for meta-analysis. Res Synth Methods [Internet]. 2010 Apr [cited 2022 Jun 7];1(2):112–25. Available from: https://pubmed.ncbi.nlm.nih.gov/26061377/

29. Begg CB, Mazumdar M. Operating Characteristics of a Rank Correlation Test for Publication Bias. Biometrics. 1994 Dec;50(4):1088.

30. Sterne JAC, Egger M. Regression methods to detect publication and other bias in meta-analysis. Publication Bias in Meta-Analysis: Prevention, Assessment and Adjustments. 2006 Jan 1;99–110.

31. Spensley KJ, Gleeson S, Martin P, Thomson T, Clarke CL, Pickard G, et al. Comparison of Vaccine Effectiveness Against the Omicron (B.1.1.529) Variant in Hemodialysis Patients. Kidney Int Rep [Internet]. 2022 Apr [cited 2022 Jun 7];7(6). Available from: https://pubmed.ncbi.nlm.nih.gov/35434428/

32. Gray MBBCH GE, Collie S, Garrett MBBS N, Goga A, Champion J, Zylstra M, et al. Vaccine effectiveness against hospital admission in South African health care workers who received a homologous booster of Ad26.COV2 during an Omicron COVID19 wave: Preliminary Results of the Sisonke 2 Study. medRxiv [Internet]. 2021 Dec 29 [cited 2022 Jun 7];2021.12.28.21268436. Available from: https://www.medrxiv.org/content/10.1101/2021.12.28.21268436v1

33. Tseng HF, Ackerson BK, Luo Y, Sy LS, Talarico CA, Tian Y, et al. Effectiveness of mRNA-1273 against SARS-CoV-2 Omicron and Delta variants. Nat Med [Internet]. 2022 May 1 [cited 2022 Jun 7];28(5). Available from: https://pubmed.ncbi.nlm.nih.gov/35189624/

34. Accorsi EK, Britton A, Fleming-Dutra KE, Smith ZR, Shang N, Derado G, et al. Association Between 3 Doses of mRNA COVID-19 Vaccine and Symptomatic Infection Caused by the SARS-CoV-2 Omicron and Delta Variants. JAMA [Internet]. 2022 Feb 15 [cited 2022 Jun 7];327(7):639–51. Available from: https://pubmed.ncbi.nlm.nih.gov/35060999/

35. Andeweg SP, Gier B de, Eggink D, Ende C van den, Maarseveen N van, Ali L, et al. Protection of COVID-19 vaccination and previous infection against Omicron BA.1, BA.2 and Delta SARS-CoV-2 infections. medRxiv [Internet]. 2022 May 12 [cited 2022 Jun 7];2022.02.06.22270457. Available from: https://www.medrxiv.org/content/10.1101/2022.02.06.22270457v3

36. Chemaitelly H, Tang P, Hasan MR, AlMukdad S, Yassine HM, Benslimane FM, et al. Waning of BNT162b2 Vaccine Protection against SARS-CoV-2 Infection in Qatar. New England Journal of Medicine [Internet]. 2021 Dec 9 [cited 2022 Jun 7];385(24):e83. Available from: https://www.nejm.org/doi/full/10.1056/NEJMoa2114114

37. Powell AA, Kirsebom F, Stowe J, McOwat K, Saliba V, Ramsay ME, et al. Adolescent vaccination with BNT162b2 (Comirnaty, Pfizer-BioNTech) vaccine and effectiveness of the first dose against COVID-19: national test-negative case-control study, England. medRxiv [Internet]. 2021 Dec 11 [cited 2022 Jun 7];2021.12.10.21267408. Available from: https://www.medrxiv.org/content/10.1101/2021.12.10.21267408v1

38. Sheikh A, Kerr S, Woolhouse M, McMenamin J, Robertson C. Severity of Omicron variant of concern and vaccine effectiveness against symptomatic disease: national cohort with nested test negative design study in Scotland. G. Balint, Antala B, Carty C, Mabieme JMA, Amar IB, Kaplanova A, editors. Uniwersytet śląski [Internet]. 2013 [cited 2022 Jun 7];343–54. Available from: https://sbc.org.pl/dlibra/publication/99008/edition/93276/synteza-i-aktywnosc-biologiczna-nowych-analogow-tiosemikarbazonowych-chelatorow-zelaza-serda-maciej?language=en

39. Collie S, Champion J, Moultrie H, Bekker LG, Gray G. Effectiveness of BNT162b2 Vaccine against Omicron Variant in South Africa. New England Journal of Medicine [Internet]. 2022 Feb 3 [cited 2022 Jun 8];386(5):494–6. Available from: https://www.nejm.org/doi/full/10.1056/NEJMc2119270

40. Ferdinands JM, Rao S, Dixon BE, Mitchell PK, DeSilva MB, Irving SA, et al. Waning 2-Dose and 3-Dose Effectiveness of mRNA Vaccines Against COVID-19–Associated Emergency Department and Urgent Care Encounters and Hospitalizations Among Adults During Periods of Delta and Omicron Variant Predominance — VISION Network, 10 States, August 2021–January 2022. MMWR Morbidity and Mortality Weekly Report [Internet]. 2022 [cited 2022 Jun 8];71(7):255–63. Available from: https://www.cdc.gov/mmwr/volumes/71/wr/mm7107e2.htm

41. Tartof SY, Slezak JM, Puzniak L, Hong V, Xie F, Ackerson BK, et al. BNT162b2 (Pfizer–Biontech) mRNA COVID-19 Vaccine Against Omicron-Related Hospital and Emergency Department Admission in a Large US Health System: A Test-Negative Design. SSRN Electronic Journal [Internet]. 2022 Jan 20 [cited 2022 Jun 8]; Available from: https://papers.ssrn.com/abstract=4011905

42. Thompson MG, Natarajan K, Irving SA, Rowley EA, Griggs EP, Gaglani M, et al. Effectiveness of a Third Dose of mRNA Vaccines Against COVID-19–Associated Emergency Department and Urgent Care Encounters and Hospitalizations Among Adults During Periods of Delta and Omicron Variant Predominance — VISION Network, 10 States, August 2021–January 2022. MMWR Morbidity and Mortality Weekly Report [Internet]. 2022 Jan 21 [cited 2022 Jun 8];71(4):139–45. Available from: https://www.cdc.gov/mmwr/volumes/71/wr/mm7104e3.htm

43. Polack FP, Thomas SJ, Kitchin N, Absalon J, Gurtman A, Lockhart S, et al. Safety and Efficacy of the BNT162b2 mRNA Covid-19 Vaccine. N Engl J Med [Internet]. 2020 Dec 31 [cited 2022 Jun 8];383(27):2603–15. Available from: https://pubmed.ncbi.nlm.nih.gov/33301246/

44. Baden LR, el Sahly HM, Essink B, Kotloff K, Frey S, Novak R, et al. Efficacy and Safety of the mRNA-1273 SARS-CoV-2 Vaccine. New England Journal of Medicine. 2021 Feb 4;384(5):403–16.

45. Sadoff J, Gray G, Vandebosch A, Cárdenas V, Shukarev G, Grinsztejn B, et al. Safety and Efficacy of Single-Dose Ad26.COV2.S Vaccine against Covid-19. New England Journal of Medicine [Internet]. 2021 Jun 10 [cited 2022 Jun 8];384(23):2187–201. Available from: https://www.nejm.org/doi/full/10.1056/NEJMoa2101544

46. Lin DY, Gu Y, Wheeler B, Young H, Holloway S, Sunny SK, et al. Effectiveness of Covid-19 Vaccines in the United States Over 9 Months: Surveillance Data from the State of North Carolina. medRxiv [Internet]. 2021 Oct 26 [cited 2022 Jun 8];2021.10.25.21265304. Available from: https://www.medrxiv.org/content/10.1101/2021.10.25.21265304v1

