## Supplementary material for "Effectiveness of vaccination against SARS-CoV-2 Omicron variant infection, symptomatic disease, and hospitalisation: a systematic review and meta-analysis"

Table S1 Search strategy

Last conducted on: 01/03/2022

| Database | Link | Query | Results |
|---|---|---|---|
| PubMed | https://pubmed.ncbi.nlm.nih.gov/?term=%28vaccine+AND+effectiveness%29+AND+%28Omicron+OR+B.1.1.529%29+AND+LitCPREVENTION%5Bfilter%5D&sort=date&size=100 | (vaccine AND effectiveness) AND (Omicron OR B.1.1.529) AND LitCPREVENTION[filter] | 73 |
| medRxiv+bioRxiv | https://www.medrxiv.org/search/%2528covid-19%252Bor%252Bsars-cov-2%2529%252Band%252Bvaccine%252Band%252Beffectiveness%252Band%252B%2528Omicron%252Bor%252BB.1.1.529%2529 | (covid-19 or sars-cov-2) and vaccine and effectiveness and (Omicron or B.1.1.529) | 412 |
| WHO COVID-19 DATABASE | https://search.bvsalud.org/global-literature-on-novel-coronavirus-2019-ncov/?output=site&lang=en&from=1&sort=&format=summary&count=20&fb=&page=1&skfp=&index=ab&q=vaccine+AND+effectiveness+AND+%28Omicron+OR+B.1.1.529%29&search_form_submit= | vaccine AND effectiveness AND (Omicron OR B.1.1.529) | 64 |
| View-Hub | https://view-hub.org/covid-19/effectiveness-studies?field_covid_studies_variant_tabl=8536 | | 24 |

Table S2. Characteristics of included studies

| | Author (publ. date) | Outcome measure | Vaccine | Country | Design | Population | Max Duration of follow up after fully vaccinated (~weeks) |
|---|---|---|---|---|---|---|---|
| 1 | Accorsi et al. (January 21, 2022) | Symptomatic infection | mRNA-1273 BNT162b2 | USA | Test-negative case control | 52,978 tests across 49 states in USA | 3 |
| 2 | Andeweg et al. (February 8, 2022) | Symptomatic infection | mRNA-1273 BNT162b2 ChAdOx1; BNT162b2 and mRNA-1273 booster | Netherlands | Test-negative case control | 528,488 tests: 448,268 negative, 41,245 (7.8%) positive for Delta and 38,974 (7.4%) positive for Omicron | 8 |
| 3 | UK HSA (January 14, 2021) (update of Andrews et al. publication) | Hospitalisation | mRNA-1273 BNT162b2 ChAdOx1 | England | Test-negative case control | 760,647 Omicron cases, 236,023 Delta cases, and test negative controls aged 18+ | 14 |
| 4 | Buchan et al. (January28,2022) | Documented infection | mRNA, booster BNT162b2 | Canada | Test negative case control | 16,087 Omicron-positive cases, 4261 Delta-positive cases, and 114,087 test-negative controls aged ≥18 years | 34 |
| 5 | Chemaitelly et al. (March 13,2022) | Symptomatic/Severe, critical or fatal disease | BNT162b2, mRNA1273 | Qatar | Test-negative case control | 138,182 individuals | 58 |
| 6 | Collie et al. (December 29, 2021) | Hospitalisation | BNT162b2 | South Africa | Test negative case control | 211,610 PCR tests of individuals In Gauteng Province | 24 |
| 7 | Ferdinands et al. (February 18, 2022) | ED or UC encounters, Hospitalisation | BNT162b2, mRNA1273 | USA | Test negative case control | 241,204 ED/UC encounters and 93,408 hospitalisations | 25 |
| 8 | Gray et al. (December 29,2021) | Hospitalisation | Ad26.COV.2 | South Africa | Test-negative case control | 69,092 HCWs | 13 |
| 9 | Powell et al. (February 11, 2022) | Symptomatic disease | BNT162b2 | UK | Test-negative case control | 61,543 eligible tests for 16-17 years olds | 33 |
| 10 | Sheikh et al. (December 22,2021) | Symptomatic disease | BNT162b2, mRNA1273, AZD1222 primary series+ BNT162b2 and mRNA-1273 booster | Scotland | Test-negative case control | 162,946 RT-PCR positive tests in Scotland | 7 |
| 11 | Spensley et al. (January 26, 2022) | Documented infection | BNT162b2 AZD1222 | UK | Prospective cohort | 1121 end stage kidney disease patients receiving in-center haemodialysis | 52 |
| 12 | Tartof et al. (January 18, 2022) | ED admission, Hospitalisation | BNT162b2 | USA | Test-negative case control | 8694 hospital admissions, and 11,719 ED admissions in Southern California | 44 |
| 13 | Thompson et al. (January 21,2022) | ED or UC encounters, Hospitalisation | BNT162b2 & mRNA-1273 | USA | Test-negative case control | 222,772 ED encounters and 87,904 hospital admissions | 32 |
| 14 | Tseng et al. (January 21, 2022) | Documented infection | mRNA-1273 | USA | Test negative case control | 75,630 Kaiser Permanent Southern California members aged 18+ | 47 |
| 15 | Willet et al. (Janaury 26,2021) | Documented infection | BNT162b2 mRNA-1273 AZD1222 | Scotland | Test-negative case control | 6166 Omicron cases and 4911 Delta cases | 11 |

**Table S3.** Characteristics of the observations included in the quantitative synthesis of the primary outcome

| Characteristics | Total (N=55) | Vaccination: Booster (N=28) | Vaccination: Primary course (N=27) |
|---|---|---|---|
| **Study design** | | | |
| case-negative control | 51 (93 %) | 26 (93 %) | 25 (93 %) |
| cohort | 4 (7 %) | 2 (7 %) | 2 (7 %) |
| **Country** | | | |
| Canada | 3 (5 %) | 2 (7 %) | 1 (4 %) |
| Netherlands | 2 (4 %) | 1 (4 %) | 1 (4 %) |
| Qatar | 8 (15 %) | 4 (14 %) | 4 (15 %) |
| South Africa | 2 (4 %) | 0 (0 %) | 2 (7 %) |
| UK | 19 (35 %) | 8 (29 %) | 11 (41 %) |
| US | 21 (38 %) | 13 (46 %) | 8 (30 %) |
| **Sample age (years)** | | | |
| Mean (SD) | 44.7 (10.4) | 44.4 (9.06) | 45.0 (11.7) |
| Median [Min, Max] | 41.0 [16.5, 74.0] | 41.0 [34.0, 65.0] | 41.0 [16.5, 74.0] |
| **Elderly** | | | |
| <65 years | 53 (96 %) | 27 (96 %) | 26 (96 %) |
| >65 years | 2 (4 %) | 1 (4 %) | 1 (4 %) |
| **Study follow-up period (days)** | | | |
| Mean (SD) | 142 (99.9) | 74.6 (41.0) | 212 (95.1) |
| Median [Min, Max] | 105 [14.0, 365] | 62.0 [14.0, 150] | 213 [70.0, 365] |
| **Routine rt-PCR test surveillance** | | | |
| rt-PCR routine surveillance | 18 (33 %) | 9 (32 %) | 9 (33 %) |
| NO rt-PCR routine surveillance | 37 (67 %) | 19 (68 %) | 18 (67 %) |
| **Induction time (days)** | | | |
| Mean (SD) | 13.3 (5.66) | 10.8 (3.56) | 15.9 (6.29) |
| Median [Min, Max] | 14.0 [7.00, 30.0] | 14.0 [7.00, 14.0] | 14.0 [7.00, 30.0] |
| **Primary vaccination course** | | | |
| Ad26.COV.2 | 2 (4 %) | 2 (7 %) | 0 (0 %) |
| BNT162b2 | 18 (33 %) | 8 (29 %) | 10 (37 %) |
| BNT162b2/mRNA-1273 | 8 (15 %) | 4 (14 %) | 4 (15 %) |
| ChAdOx1-S | 5 (9 %) | 2 (7 %) | 3 (11 %) |
| ChAdOx1-S/BNT16b2/mRNA-1273 | 7 (13 %) | 4 (14 %) | 3 (11 %) |
| mRNA-1273 | 15 (27 %) | 8 (29 %) | 7 (26 %) |
| **Primary vaccination: vaccine technology** | | | |
| mRNA | 41 (75 %) | 20 (71 %) | 21 (78 %) |
| VV | 6 (11 %) | 4 (14 %) | 2 (7 %) |
| VV/mRNA | 8 (15 %) | 4 (14 %) | 4 (15 %) |
| **Booster: vaccine technology** | | | |
| mRNA | 26 (47 %) | 26 (93 %) | 0 (0 %) |
| VV | 2 (4 %) | 2 (7 %) | 0 (0 %) |
| | 0 (0 %) | 0 (0 %) | 0 (0 %) |
| **Clinical severity of Omicron infection** | | | |
| Symptomatic | 24 (44 %) | 12 (43 %) | 12 (44 %) |
| Hospitalized | 14 (25 %) | 7 (25 %) | 7 (26 %) |
| Any positive PCR | 17 (31 %) | 9 (32 %) | 8 (30 %) |
| **Risk of bias (Robins)** | | | |
| Low | 10 (18 %) | 6 (21 %) | 4 (15 %) |
| Moderate | 17 (31 %) | 6 (21 %) | 11 (41 %) |
| Serious | 28 (51 %) | 16 (57 %) | 12 (44 %) |

### 1. Analysis of heterogeneity

#### 1.1 Primary course vaccination: effectiveness against Omicron VOC

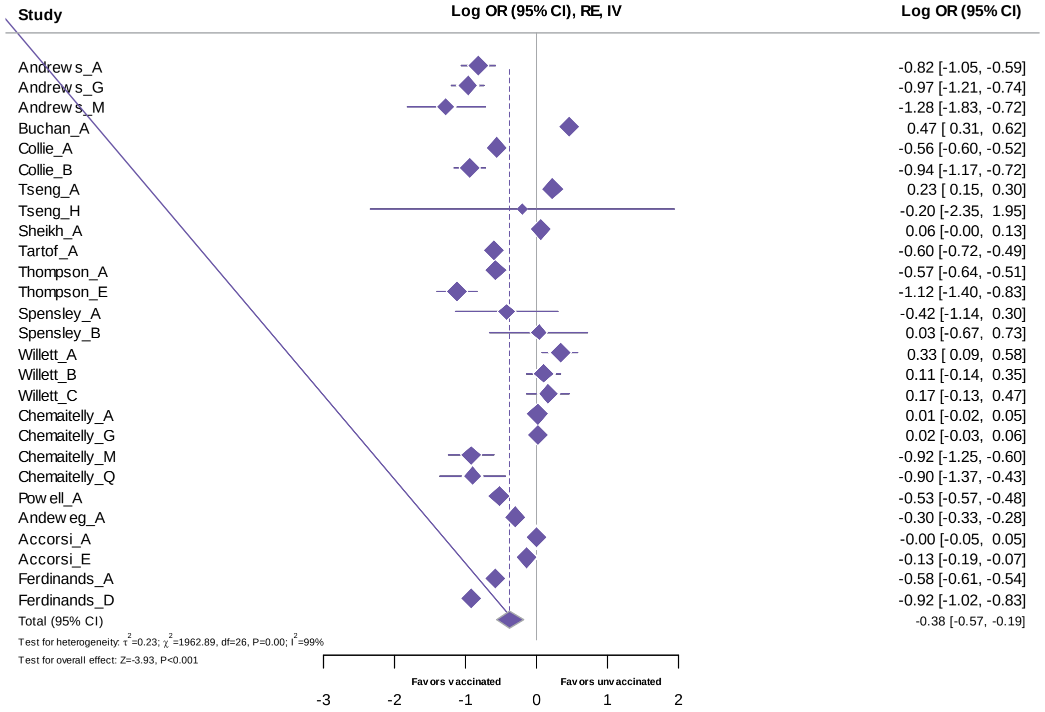

**Figure S1. Forest plot.** Results of 27 observations examining the overall risk of Sars-Cov2 infection (B.1.1.529 VOC) after a primary course vaccination. Meta-analysis based on Random Effects model, inverse variance method (IV). Effect size estimates expressed in Log Odds Ratio [95%CI].

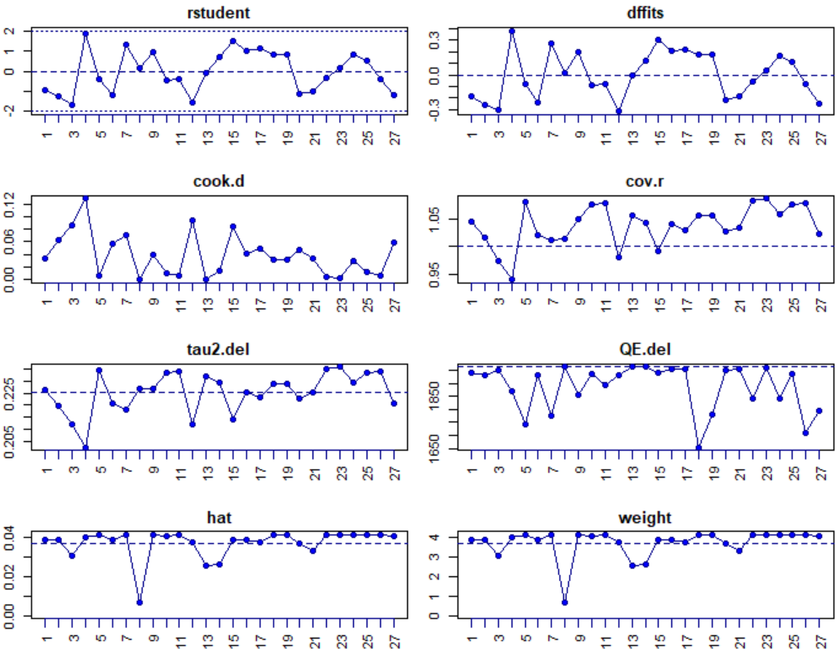

**Figure S2. Outliers and influential case diagnostic.** The influence analysis did not detect any potentially influential study because the output did not mark with an asterisk (*) any of them. Studentized residuals and Cook's distances are used to examine whether studies may be outliers and/or influential in the context of the model. Studies with a studentized residual larger than the 100×(1-0.05/(2×k))100×(1-0.05/(2×k))$^{th}$ percentile of a standard normal distribution are considered potential outliers (i.e., using a Bonferroni correction with two-sided α=0.05 for k studies included in the meta-analysis). The normality assumption is evaluated via QQ normal plot. (Viechtbauer & Cheung, 2010) When plotted, effect sizes/studies determined to be influential cases according to the rules described in Viechtbauer & Cheung (2010) are shown in red. In particular, the studentized residuals do not deviate noticeably from the model in any study suggesting a potential outlier. The DFFITS value indicates how many standard deviations the predicted (average) effect for the i$^{th}$ case changes after excluding that case from the model fitting. Cook's distance is interpreted as the Mahalanobis distance between the entire set of predicted values once with the i$^{th}$ case included and once with the i$^{th}$ case excluded from the model fitting. In this case, none of the studies could be considered to be overly influential.

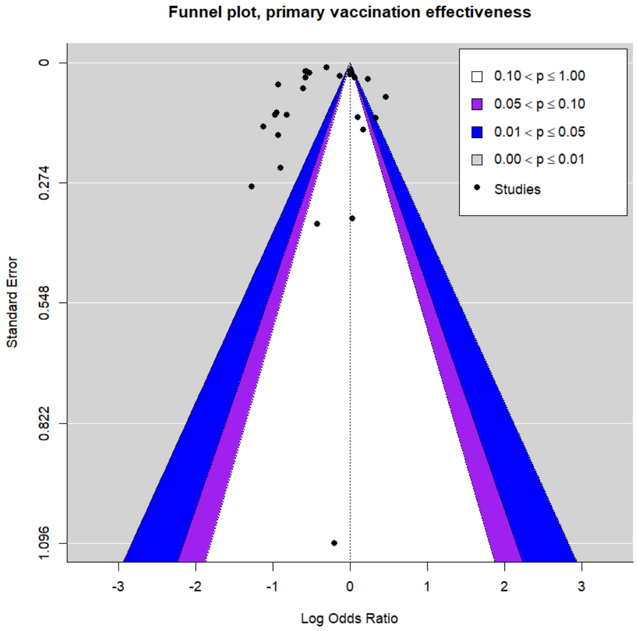

**Figure S3. Funnel plots, primary vaccination effectiveness against Sars-Cov2 (B.1.1.529 VOC).** The funnel plots are centered at 0 (i.e., at the value under the null hypothesis of no effect) and display the studies' results (x-axis) and their precision (y-axis). In both the meta-analyses, the results are expressed as log odds ratios (ORs) and the precision is represented by the standard error of the estimates. Each dot represents a single observation. The three levels of statistical significance of the effect sizes correspond by the shaded regions. If studies appear to be missing in areas of low statistical significance, then it is possible that the asymmetry is attributable to publication bias. Conversely, if studies are missing in the areas of high statistical significance, the publication bias is less likely because of the funnel asymmetry (Peter et al., 2010) The funnel plot does not show clear asymmetry and the asymmetry tests, including the Egger's test, are not significant (assuming a level of significance at 0.05).

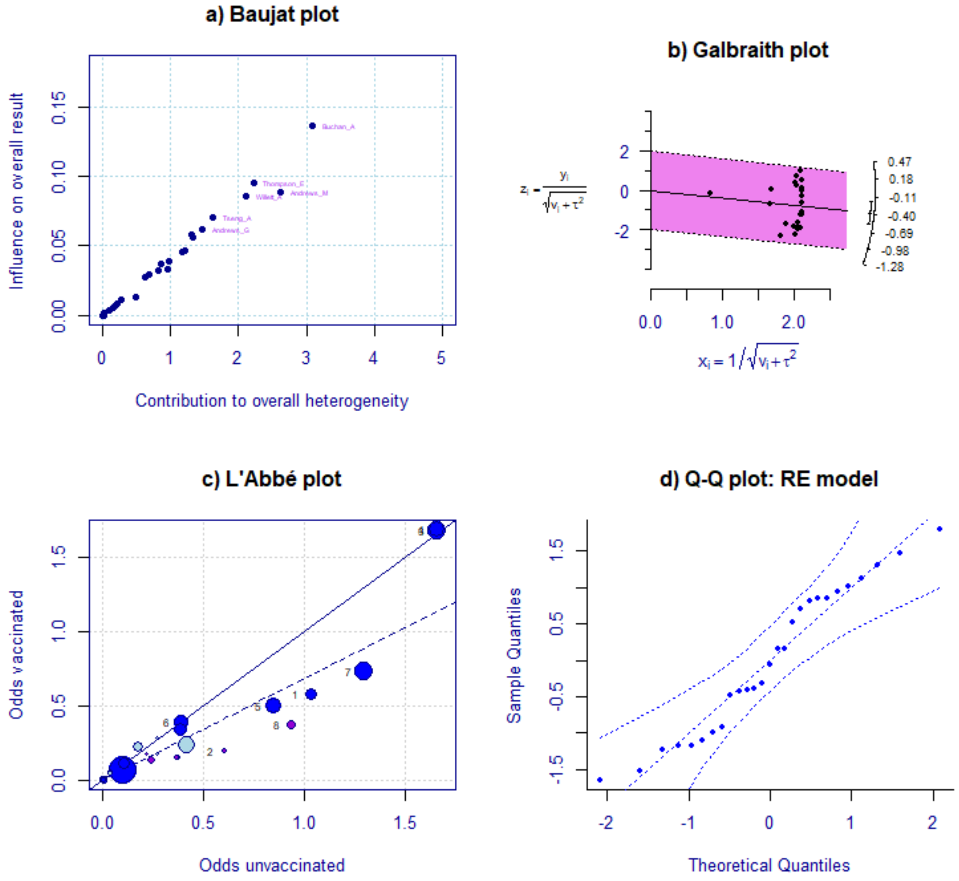

**Figure S4. Baujat plot (a)** The Baujat plot evaluates heterogeneity patterns in the meta-analysis (Baujat et al., 2002). The x-axis of the Baujat plot shows the overall heterogeneity contribution of each effect size (study) while the y-axis shows the influence of each effect size on the pooled result. Effect sizes or studies with high values on both the x and y-axis may be considered influential cases, such as Buchan_A. **Galbraith (radial) plot (b).** The radial plot represents a way to assess the consistency of observed outcomes with differing precisions (attributable to heteroscedastic sampling variances). For a random effects model, the function applies $1/\sqrt{v_i + \tau^2}$ for the horizontal and $y_i/\sqrt{v_i + \tau^2}$ for the vertical axis. On the right hand side of the plot, the arc indicates the value of the observed outcome for that point while the central line indicates the pooled effect with the slop equal to the pooled effect Log Odds= −0.38 (95% CI: −0.57 to −0.19). (Galbraith, 1988) This plot does not show for outliers in the effect sizes because the 95% of the studies lay within the area defined by the two (lighter coloured) confidence interval lines. **L'Abbé plot (c)** The L'Abbé plot allows inspection of the variability between experimental (vaccinated) and control (unvaccinated) group outcomes. (L'Abbé, Detsky, & O'Rourke, 1987) The dashed line indicates the overall estimate based on the fitted model (which is linear on the log scale for the log odds ratio). The solid diagonal line represents the indifference effect line. For the points laying on the diagonal line, the risk of infection did not differ between the two groups. For the points below this line, the risk was lower in the vaccinated group. The size of the points is an inverse function of the precision of the estimates. The colour of the dots indicate different infection severity risk (light =any infection; medium=symptomatic; dark=hospitalization) The largest symptomatic infection estimates lay on the indifference line while the effect sizes corresponding to hospitalization are below the average pooled effect size. **Q-Q normal plot (d)** In this plot the theoretical quantiles of a normal distribution are plotted on the horizontal axis against the observed quantiles of the studentized residuals (vertical axis). In the QQ normal plot, the points should ideally fall on a diagonal line with slope of 1, going through the (0, 0) point. Any deviation from the X = Y reveals how the distributions differ from the normal. Deviations from the diagonal may indicate that the residual heterogeneity in the true effects is non-normally distributed as well as there are subgroups in the data, or that publication bias might present. (Wang & Bushman, 1998) The pseudo confidence envelope is created based on the quantiles of 1000 simulated sets of pseudo residuals from the model. The simulated bounds are smoothed with Friedman's Super-Smoother. (Luedicke, 2015) Although not every observation fall on the diagonal line, no study falls outside the pseudo-confidence envelope.

`

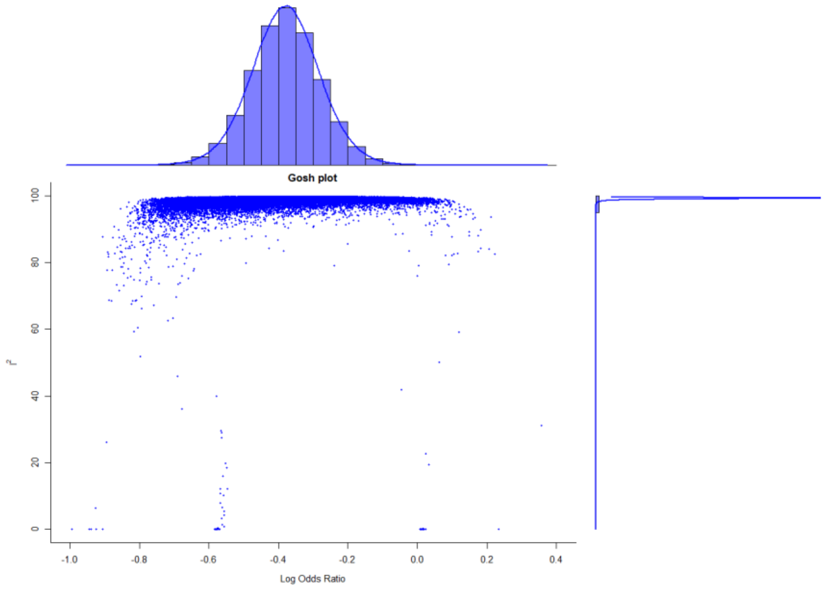

**Figure S5. GOSH (Graphic Display of Heterogeneity) plot. Effectiveness of primary vaccination meta-analysis.** The analysis is based on examining the results of an equal-effects model in all possible subsets of the n=27 studies included in the meta-analysis. In a homogeneous set of studies, the model estimates form a roughly symmetric, contiguous, and unimodal distribution such as the bell curve obtained in this case. Conversely, a multimodal distribution may suggests the presence of heterogeneity, possibly due to outliers and/or distinct subgroupings of studies. By plotting the estimates against some measure of heterogeneity (e.g., $I^2$) it may detect sub clusters, which are indicative of heterogeneity.

#### 1.2 *One booster dose vaccination: effectiveness against Omicron VOC*

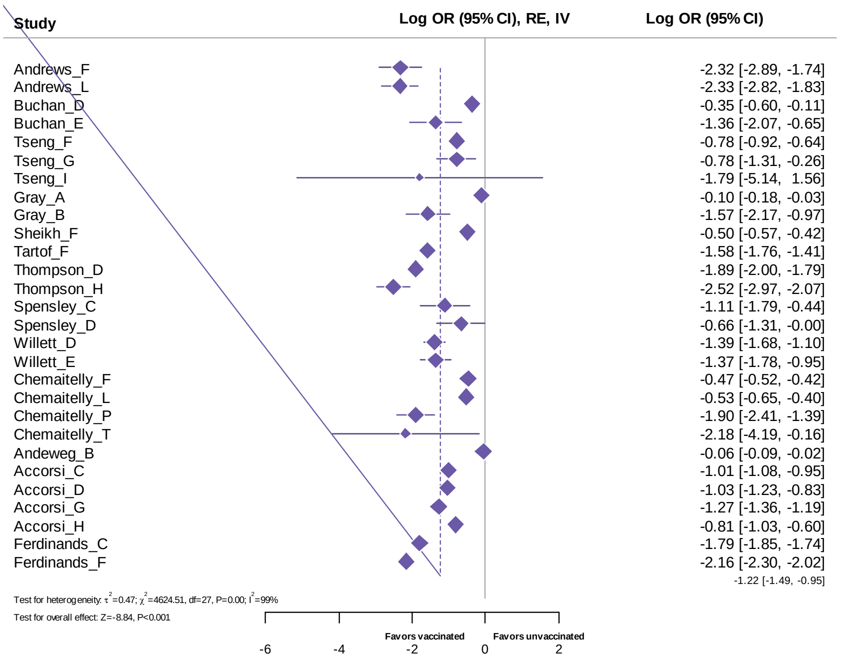

**Figure S6. Forest plot.** Results of 28 observations examining the overall risk of Sars-Cov2 infection (B.1.1.529 VOC) after one booster dose. Meta-analysis based on Random Effects (RE) model, inverse variance method (IV). Effect size estimates expressed in Log Odds Ratio [95%CI].

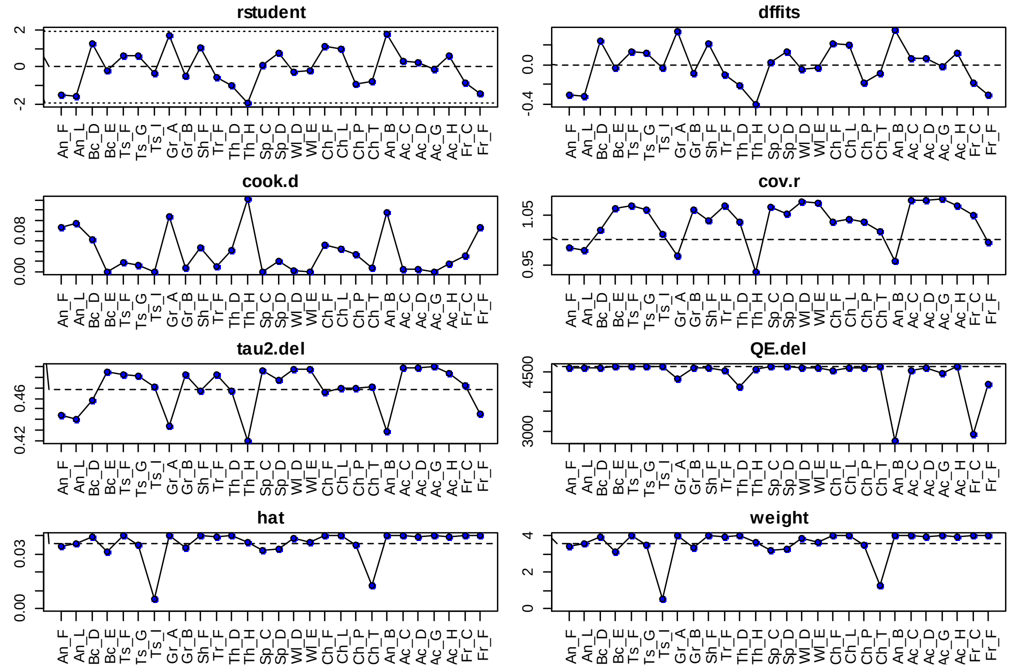

Figure S7. Outliers and influential case diagnostic. Booster dose effectiveness. According to the Q-test, the true outcomes appear to be heterogeneous (Q (27) =4624.51, $p<0.0001$; $\tau^2=0.4686$; $I^2=99.33\%$). A 95% prediction interval for the true outcomes is given by −2.5843 to 0.1529. Studies with a Cook's distance larger than the median plus six times the interquartile range of the Cook's distances are considered as influential. Given the Cook's distances, none of the studies can be considered particularly influential. The studentized residuals do not deviate noticeably from the model in any observation, excluding potential outlier. The influence analysis does not detect any overly influential study.

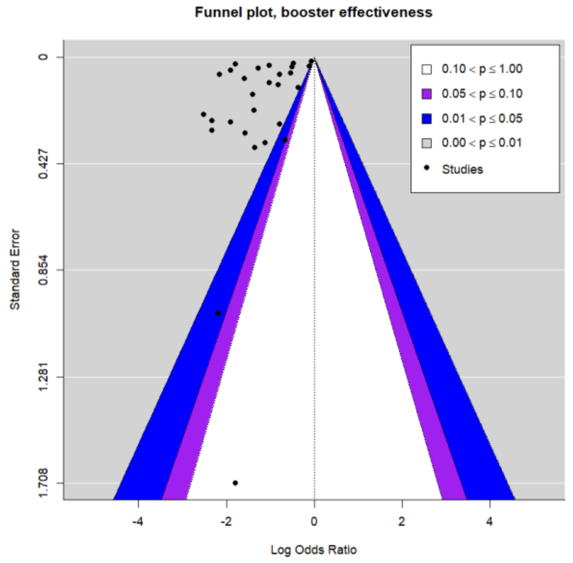

Figure S8. Funnel plots, booster effectiveness against Sars-Cov2 (B.1.1.529 VOC). The funnel plots are centered at 0 (i.e., at the value under the null hypothesis of no effect) and display the studies' results (x-axis) and their precision (y-axis). In both the meta-analyses, the results are expressed as log odds ratios (ORs) and the precision is represented by the standard error of the estimates. Each dot represents a single observation. The three levels of statistical significance of the effect sizes correspond by the shaded regions. Ideally, all the included studies should scatter either side of the overall effect symmetrically. (Peter et al., 2010) The observations cluster around the top, with only one observation located at the bottom, in the white region. Although the asymmetry suggests a publication bias likelihood, the asymmetry tests, including the Egger's test, are not significant (assuming a level of significance at 0.05).

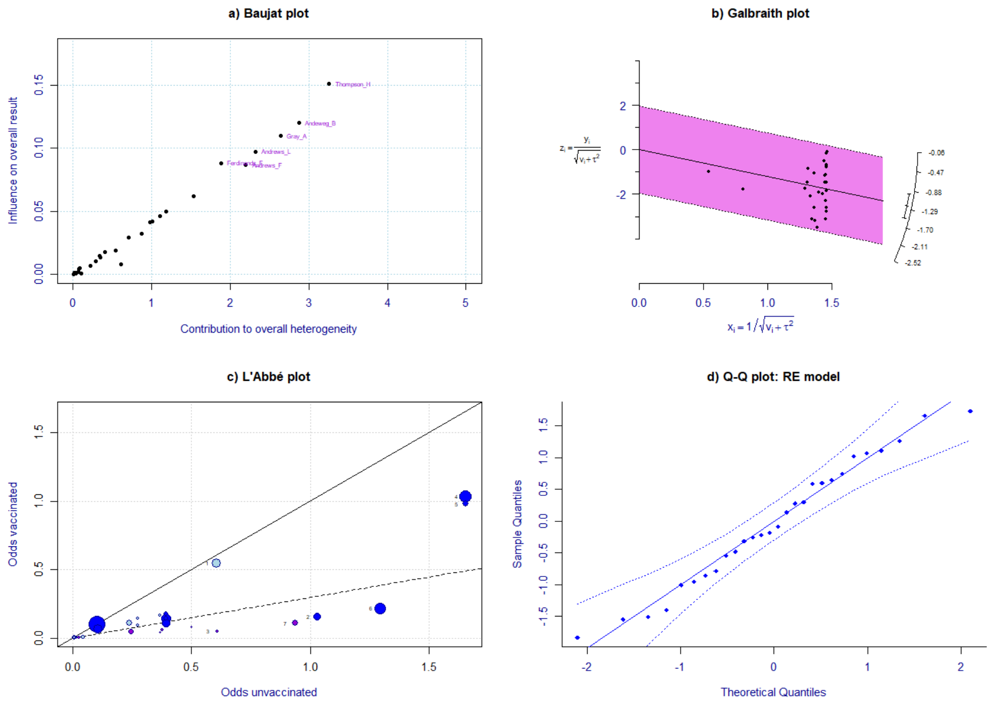

**Figure S9. Analysis of heterogeneity, effectiveness of one booster dose. Baujat plot (a)** The Baujat plot shows the contribution of each study to the overall heterogeneity (as measured by Cochran's Q) on the horizontal axis, and its influence on the pooled effect size on the vertical axis. Studies on the right side of the plot can be considered as potentially influent cases since they contribute heavily to the overall heterogeneity. (Baujat et al. 2002) The study 'Thompson_H' shows a potential influence on both the overall heterogeneity (x-axis) and on the pooled effect size (y-axis). However, none of the studies could be considered particularly influential according to the influence analysis. **Galbraith (radial) plot (b).** The Galbraith plot is a graphical option to address heterogeneity. The plot summarizes meta-analysis output with the centre line indicating the pooled effect with the slop equal to the pooled effect Log Odds= −1.22 (95%CI: −1.49 to −0.95). The plot reports information about the study-specific effect sizes and their precisions, reports the overall effect size, and helps detect potential outliers. In the absence of substantial heterogeneity, around 95% of the studies lie within the 95% CI region (shaded area). The majority of the studies are have smaller SE and larger 1/SE; hence, they aggregate away from the origin. The plot does not detect outliers in the effect sizes because the 95% of the studies lay within the area defined by the two (lighter coloured) confidence interval lines. (Galbraith, 1988) **L'Abbé plot (c)** The L'Abbé plot is a scatter plot of the log odds in the control group (x-axis) against those in the treatment group (y-axis). The between-study heterogeneity is explored by comparing group-level outcome measures across studies and by identifying outlying studies. The dashed line indicates the overall estimate based on the fitted model. (L'Abbe, Detsky, and O'Rourke, 1987) The colours represent three different levels of severity: any positive rtPCR (light), symptomatic infection (medium) and severe/hospitalization (dark). For the points laying on the diagonal line, the risk of infection did not differ between the two groups. For the points below this line, the risk was lower in the vaccinated group. The bigger points correspond to more precise estimates and *vice versa*, most of them pertains to symptomatic risk estimates. All the effect sizes fall below the pooled effect size line, except *'Andeweg_B'* on symptomatic risk which is the nearest and *'Tseng_I'* on hospitalization risk, which overlaps the indifference line. The colour of the bubbles represents three different categories of risk (light =any infection; medium=symptomatic; dark=hospitalization). The hospitalization risk estimates tend to locate below the dotted line, while the symptomatic risk effect estimates appear more scattered. **Q-Q normal plot (d)** The QQ normal plot can be used as diagnostic tool. In the graph, the theoretical quantiles of a normal distribution are plotted on the horizontal axis against the observed quantiles of the (externally) standardized residuals on the vertical axis. The deviations of the points from the diagonal line may indicate residual heterogeneity in the true effects (non-normally distributed), presence of subgroups in the data (that are not adequately modelled by the moderators included in the model), and/or publication bias. (Wang and Bushman, 1998) No study falls outside the pseudo-confidence envelope and all points are distributed nearby the diagonal line.

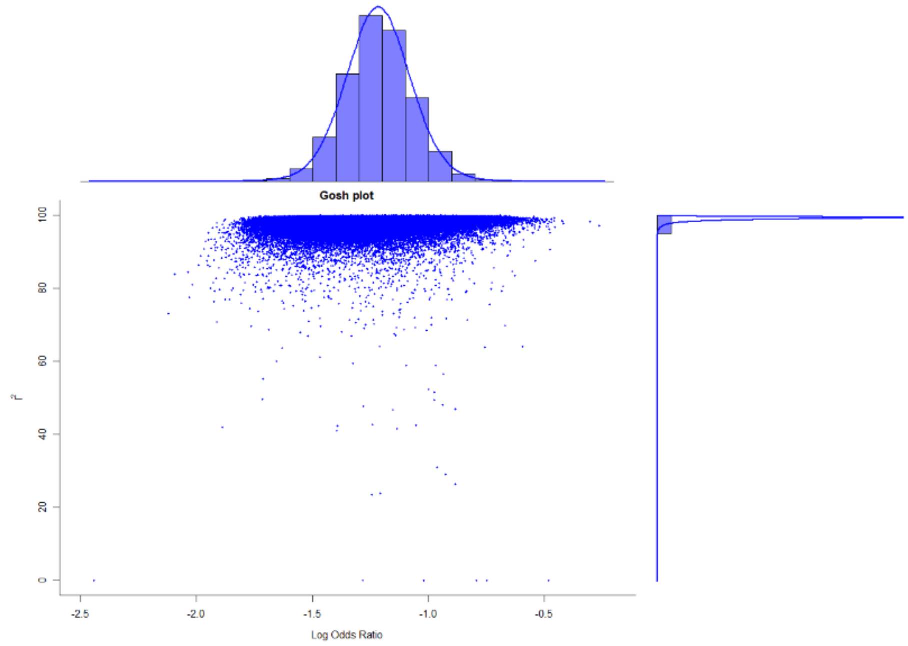

Figure S10. Graphic Display of Heterogeneity (GOSH) plots. Effectiveness of one booster dose meta-analysis. The same meta-analysis model is fitted to all possible subsets of the (n=28) included studies. Once the models are calculated, the GOSH plot displays the pooled effect size on the x-axis and the between-study heterogeneity on the y-axis. This allows detecting specific patterns, for instance clusters with different effect sizes and amounts of heterogeneity. Plotting the estimates against some measure of heterogeneity (e.g., $I^2$) may particularly detect sub clusters, which are indicative of heterogeneity. (Olkin, Dahabreh, and Trikalinos 2012) The plot is based on examining the results of an equal-effects model in all possible subsets of the n=28 observations included in the meta-analysis. The output shows a homogeneous set of studies and the model estimates form a roughly symmetric, contiguous, and unimodal distribution (bell curve).

### 2. *Subgroup analysis*

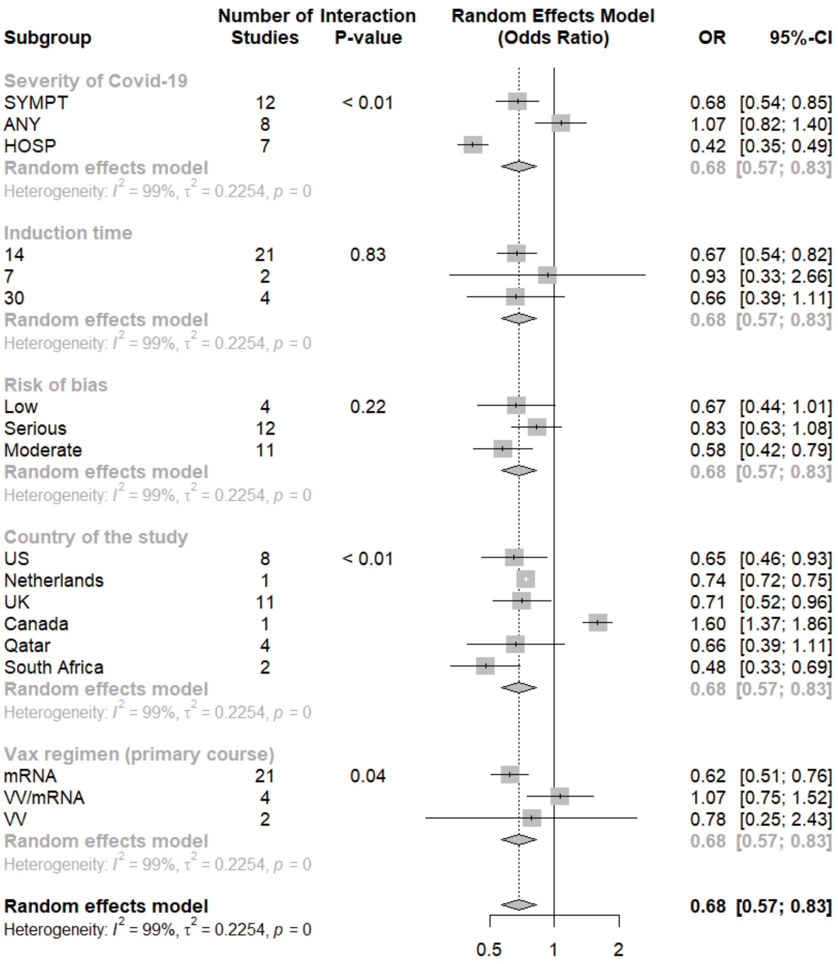

**Figure S11. Subgroup analysis for primary course vaccination effectiveness against Omicron VOC.** Random Effect model, IV method, OR [95%CI]. The subgroup analysis produces significant results for three out of five subgroups. Subgroup analyses on immunization time since the last dose (induction period) and risk of bias are not significant. In particular, studies with low risk of bias according to ROBINS-I tool, estimate a 33% reduction of risk (OR=0.67; 95%CI: 0.44 to 1.01). Studies with moderate risk of bias show a significant lower OR with risk reduction of nearly 42% (OR=0.58; 95%CI: 0.42 to 0.79), whilst studies with serious risk of bias do not produce significant OR (0.83; 95%CI=0.63-1.08). Finally, the subgroup analysis by country generates similar estimates for studies conducted in US and UK (nearly 33% and 35% risk reduction, respectively), while studies conducted in Qatar show a risk reduction of about 19% (OR=0.81; 95%CI=0.69-0.95). Regarding the vaccines used for the primary vaccination, estimates are significant for the messenger RNA (mRNA) vaccine only, which exhibits a risk reduction of 38% compared to unvaccinated (OR=0.62; 95%CI: 0.51 to 0.76). The difference in effectiveness between the homologous vaccine regimens compared to the heterologous vaccination is unclear because only observation on the messenger RNA (mRNA) vaccine exhibits a significant results, with an average risk reduction of 38% compared to unvaccinated (OR=0.62; 95%CI: 0.51 to 0.76).

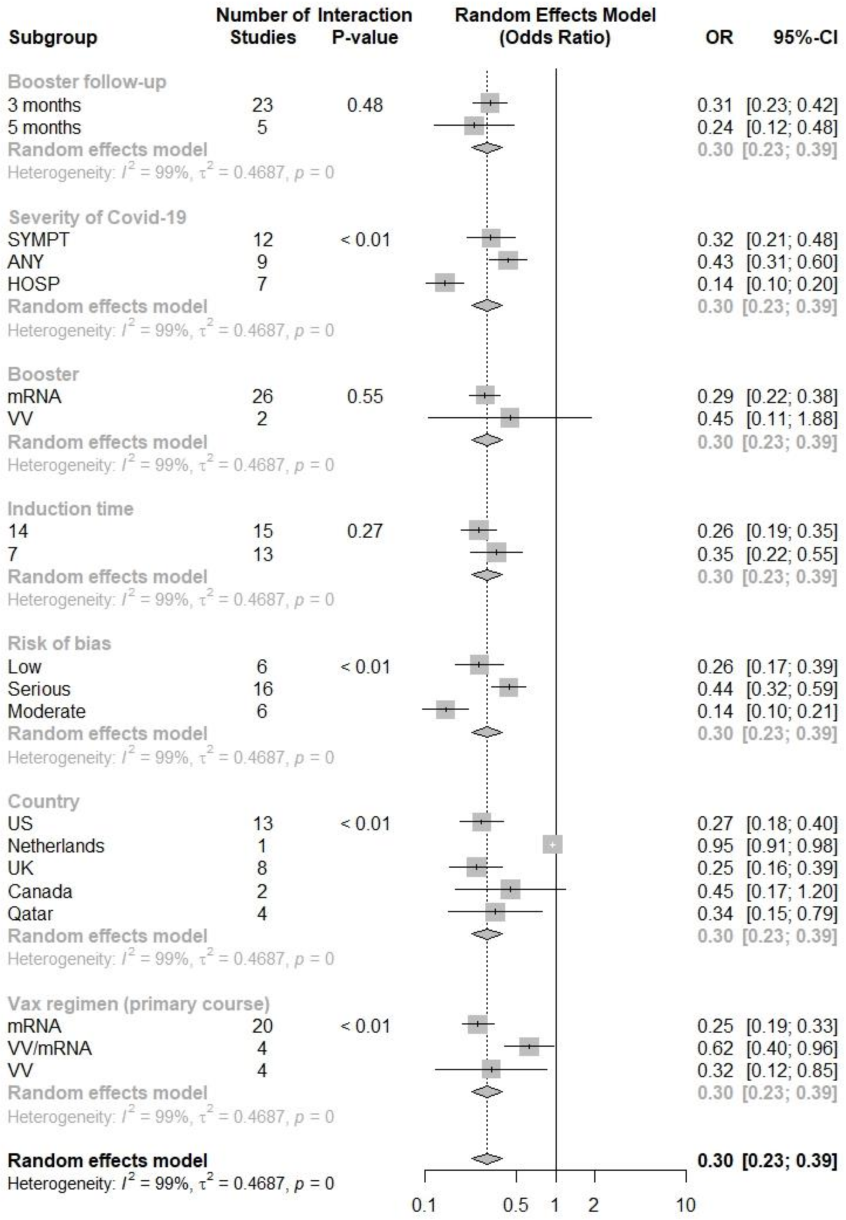

**Figure S12. Subgroup analysis for one booster effectiveness against Omicron VOC.** Random Effect model, IV method, OR [95%CI]. Random Effect model, IV method, OR [95%CI]. The subgroup analysis produces significant results for four out of six subgroups. The risk reduction for the booster group is 69% for vaccinated in studies reporting 3 months of follow up (OR= 0.31; 95%CI: 0.23 to 0.42) and 76% in studies reporting 5 months follow-up (OR= 0.24; 95%CI: 0.12 to 0.48). However, the interaction is not significant. Subgroup analyses on immunization time (induction period) and type of booster vaccine are not significant. The booster is more effective against hospitalization and symptomatic infection, yielding a 86% (OR=0.14; 95%CI: 0.10 to 0.20) and 68% (OR=0.32; 95%CI: 0.21 to 0.48) risk reduction with respect to unvaccinated respectively, while it appears less effective against any positive rt-PCR with 57% risk reduction (OR=0.43; 95%CI: 0.31 to 0.60). Studies with low risk of bias according to ROBINS-I tool, estimate a smaller OR (OR=0.26; 95%CI: 0.17 to 0.39) than studies with moderate risk of bias, while studies with serious risk of bias estimate a lower OR than studies with low risk of bias. The subgroup analysis by country generates similar estimates for studies conducted in US and UK (nearly 73% and 75% risk reduction, respectively), while studies conducted in Qatar show a risk reduction of about 66% (OR=0.34; 95%CI=0.15 to 0.79). Regarding the vaccines used for the primary vaccination (regardless the booster dose), all estimates are significant. The messenger RNA (mRNA) primary vaccination seems more protective than viral vector (VV) (OR=0.32; 95%CI: 0.12 to 0.85) and heterologous vaccination (OR=0.62; 95%CI: 0.40 to 0.96), showing a risk reduction of 75% compared to unvaccinated (OR=0.25; 95%CI: 0.19 to 0.33).

### 3. Meta-regression

#### 3.1 Primary outcomes

**Primary course vaccination**

**a) Random-Effects Model (k = 27; tau^2 estimator: REML)**

| logLik | deviance | AIC | BIC | AICc |
|---|---|---|---|---|
| -19.1654 | 38.3308 | 42.3308 | 44.8470 | 42.8525 |

**tau^2 (estimated amount of total heterogeneity): 0.2254 (SE = 0.0683)**
tau (square root of estimated tau^2 value): 0.4748
I^2 (total heterogeneity / total variability): 99.49%
H^2 (total variability / sampling variability): 194.30

**Test for Heterogeneity:**
Q(df = 26) = 1962.8944, p-val < .0001

**Model Results:**

| estimate | se | zval | pval | ci.lb | ci.ub | |
|---|---|---|---|---|---|---|
| -0.3788 | 0.0964 | -3.9309 | <.0001 | -0.5677 | -0.1899 | *** |

**b) Mixed-Effects Model (k = 27; tau^2 estimator: REML)**

| logLik | deviance | AIC | BIC | AICc |
|---|---|---|---|---|
| -10.6323 | 21.2647 | 29.2647 | 33.9769 | 31.3699 |

**tau^2 (estimated amount of residual heterogeneity): 0.1136 (SE = 0.0375)**
tau (square root of estimated tau^2 value): 0.3370
I^2 (residual heterogeneity / unaccounted variability): 98.93%
H^2 (unaccounted variability / sampling variability): 93.77
R^2 (amount of heterogeneity accounted for): 49.61%

**Test for Residual Heterogeneity:**
QE(df = 24) = 1677.7466, p-val < .0001

**Test of Moderators (coefficients 2:3):**
QM(df = 2) = 23.3031, p-val < .0001

**Model Results:**

| | estimate | se | zval | pval | ci.lb | ci.ub | |
|---|---|---|---|---|---|---|---|
| intrcpt | 0.0713 | 0.1327 | 0.5373 | 0.5910 | -0.1888 | 0.3315 | |
| factor(severity)HOSP | -0.9558 | 0.1982 | -4.8234 | <.0001 | -1.3442 | -0.5674 | *** |
| factor(severity)SYMPT | -0.4561 | 0.1663 | -2.7423 | 0.0061 | -0.7821 | -0.1301 | ** |

**c) Mixed-Effects Model (k = 27; tau^2 estimator: REML)**

| logLik | deviance | AIC | BIC | AICc |
|---|---|---|---|---|
| -5.2183 | 10.4366 | 28.4366 | 36.9366 | 48.4366 |

**tau^2 (estimated amount of residual heterogeneity): 0.0701 (SE = 0.0277)**
tau (square root of estimated tau^2 value): 0.2647
I^2 (residual heterogeneity / unaccounted variability): 97.25%
H^2 (unaccounted variability / sampling variability): 36.40

**Test for Residual Heterogeneity:**
QE(df = 19) = 448.5978, p-val < .0001

**Test of Moderators (coefficients 1:8):**
QM(df = 8) = 92.6692, p-val < .0001

**Model Results:**

| | estimate | se | zval | pval | ci.lb | ci.ub | |
|---|---|---|---|---|---|---|---|
| factor(severity)ANY | 0.2642 | 0.1921 | 1.3751 | 0.1691 | -0.1124 | 0.6408 | |
| factor(severity)HOSP | -0.7000 | 0.1894 | -3.6964 | 0.0002 | -1.0711 | -0.3288 | *** |
| factor(severity)SYMPT | -0.2807 | 0.1385 | -2.0271 | 0.0427 | -0.5522 | -0.0093 | * |
| factor(bias)Moderate | -0.4403 | 0.1767 | -2.4914 | 0.0127 | -0.7867 | -0.0939 | * |
| factor(bias)Serious | 0.0403 | 0.1826 | 0.2209 | 0.8252 | -0.3176 | 0.3983 | |
| age_c | -0.0075 | 0.0056 | -1.3365 | 0.1814 | -0.0186 | 0.0035 | |
| factor(vax)VV | 0.1660 | 0.2304 | 0.7203 | 0.4714 | -0.2857 | 0.6176 | |
| factor(vax)VV/mRNA | 0.0827 | 0.1941 | 0.4259 | 0.6702 | -0.2977 | 0.4631 | |

**Signif. codes: 0 '***' 0.001 '**' 0.01 '*' 0.05 '.' 0.1 ' ' 1**

Figure S13. Meta-regression outputs. Meta-analysis on the overall risk of Omicron infection after primary course vaccination. a) Random effect (RE) model. The overall effect is expressed as Log odds ratio and 95%CI ($\hat{\mu}$= -0.3788 (95%CI: -0.5677 to -0.1899), that corresponds to OR= 0.685 (95%CI: 0.567 to 0.827) by exponentiation. The amount of heterogeneity in the baseline RE model is estimated to be $tau^2$ = 0.2254. **Multi Level Meta-analysis** in order to explore 'severity' of Covid-19 as a potential source of heterogeneity, the effect sizes are clustered by severity of symptoms. Therefore, the variable 'author' defines the second level of the three-level model, while 'severity' defines the third level of the three-level model. The data set includes 27 effect sizes pertaining to 3 clusters of clinical severity. The same random effect is assigned to effect sizes within the same group, for both grouping variables, whereas different random effects are assigned to effect sizes having different grouping variable. In the MLM output sigma2.1 = 0.114 represents the within-group variance and sigma2.2= 0.205 the estimated value for the variance between groups. The test for heterogeneity shows signicant variation between all effect sizes in the data set, since the p value is smaller than 0.001 (Q(df = 26) = 1962.894, p < 0.001). Both the within-group variance and the between-group variance are significant at likelihood-ratio-tests (p< 0.0001 and p=0.0015, respectively). Overall, 0.36% of the overall variance can be attributed to level 1, 35.6% to level 2 ('within-group variance'), and as much as 64.1% to level 3 ('between-group variance'). **b) Mixed effect meta-regression output (one moderator).** By including the variable 'severity' as a predictor/moderator in the meta-regression model (with intercept), 'Any positive rt-PCR' factor is considered as the reference level, the model intercept represents the estimated (average) log risk ratio for 'Any positive rt-PCR. The remaining coefficients indicate how much the estimated (average) log risk ratios diverge from the intercept. Both coefficients are significantly different from zero (p>0.05). The test of moderators is significant ($Q_M$ (df =2)=23.3031, p< 0.0001), hence, the results indicate that we can reject the null hypothesis that the three-level factor is not significant. Therefore, if $\hat{\beta}_0 = 0.0713$ is the estimated average log odds ratio for observations related to 'Any positive rt-PCR' , the average log odds ratio for 'hospitalization' will be $\hat{\beta}_0 + \hat{\beta}_1 = 0.0713 - 0.9558 = -0.8845$, whereas for 'symptomatic' it will be $\hat{\beta}_3 = 0.0713 - 0.4561 = -0.3848$. By exponentiation, the coefficient for hospitalization yields an OR=0.41 (95%CI: 0.31 to 0.55) and a risk reduction of 59% with respect to unvaccinated, while the coefficient for symptomatic infection yields an OR=0.68 (95%CI: 0.56 to 0.83) and a risk reduction of 32% with respect to unvaccinated. **c) Mixed effect multiple meta-regression output (four moderators).** The estimated amount of residual heterogeneity is equal to tau2=0.0701, suggesting that (0.2254 -0.0701)/ 0.2254 = 68.9% of the total amount of heterogeneity can be explained by including four moderators in the model. The test of Moderators is significant [$Q_M$(df = 8) = 92.6692, p < 0.0001] (intercept suppressed) as well as the test for residual heterogeneity (QE(df = 19)=448.5978, p< 0.0001; $I^2$=97.25%). The variable 'age_c' corresponds to the variable 'age' centered around the mean of all the observations (44.96 years). The coefficients 'age_c' and 'vax' (indicating the primary course regimen) are not significant at p<0.05. However, the coefficient representing the mean age of the samples shows a negative association $\hat{\beta}_6 = -0.0075$ with the dependent variable (OR=0.993; 95%CI: 0.983 to 1.004). All the significant coefficients show a negative association with the dependent variable, hence, the primary vaccination is protective, in particular against hospitalization. The observations with moderate risk of bias estimate a 36% lower risk for vaccinated ($\hat{\beta}_4 = -0.4403$; OR= 0.64; 95%CI: 0.46 to 0.91) with respect to studies with low risk of bias (reference).

#### One booster dose

**a) Random-Effects Model (k = 28; tau^2 estimator: REML)**

```
logLik  deviance       AIC       BIC      AICc
-29.8699   59.7399   63.7399   66.3315   64.2399
tau^2 (estimated amount of total heterogeneity): 0.4686 (SE = 0.1410)
tau (square root of estimated tau^2 value):      0.6846
I^2 (total heterogeneity / total variability):   99.33%
H^2 (total variability / sampling variability):  148.42
Test for Heterogeneity:
Q(df = 27) = 4624.5092, p-val < .0001
Model Results:
estimate      se      zval     pval     ci.lb     ci.ub
-1.2157  0.1376  -8.8351  <.0001  -1.4854  -0.9460  ***
```

**b) Mixed-Effects Model (k = 28; tau^2 estimator: REML)**

```
logLik  deviance       AIC       BIC      AICc
-24.0492   48.0983   56.0983   60.9738   58.0983
tau^2 (estimated amount of residual heterogeneity):     0.3438 (SE = 0.1099)
tau (square root of estimated tau^2 value):             0.5863
I^2 (residual heterogeneity / unaccounted variability): 99.06%
H^2 (unaccounted variability / sampling variability):   106.16
R^2 (amount of heterogeneity accounted for):            26.64%
Test for Residual Heterogeneity:
QE(df = 25) = 3911.0446, p-val < .0001
Test of Moderators (coefficients 2:3):
QM(df = 2) = 10.8779, p-val = 0.0043
Model Results:
                             estimate      se      zval     pval     ci.lb     ci.ub
intrcpt                       -0.8551  0.2098  -4.0767  <.0001  -1.2662  -0.4440  ***
factor(severity)HOSP          -1.1030  0.3400  -3.2444  0.0012  -1.7693  -0.4367   **
factor(severity)SYMPT         -0.2783  0.2716  -1.0244  0.3057  -0.8107   0.2541
```

**c) Mixed-Effects Model (k = 28; tau^2 estimator: REML)**

```
logLik  deviance       AIC       BIC      AICc
-10.6494   21.2988   39.2988   48.2604   57.2988
tau^2 (estimated amount of residual heterogeneity):     0.1037 (SE = 0.0423)
tau (square root of estimated tau^2 value):             0.3220
I^2 (residual heterogeneity / unaccounted variability): 94.54%
H^2 (unaccounted variability / sampling variability):   18.32
Test for Residual Heterogeneity:
QE(df = 20) = 306.8591, p-val < .0001
Test of Moderators (coefficients 1:8):
QM(df = 8) = 350.4571, p-val < .0001
Model Results:
                             estimate      se      zval     pval     ci.lb     ci.ub
factor(severity)ANY           -1.1005  0.2205  -4.9918  <.0001  -1.5326  -0.6684  ***
```

| | | | | | | | |
|---|---|---|---|---|---|---|---|
| factor(severity)HOSP | -2.1144 | 0.2199 | -9.6153 | <.0001 | -2.5454 | -1.6834 | *** |
| factor(severity)SYMPT | -1.1845 | 0.1391 | -8.5164 | <.0001 | -1.4571 | -0.9119 | *** |
| factor(bias)Moderate | -0.7112 | 0.2293 | -3.1011 | 0.0019 | -1.1607 | -0.2617 | ** |
| factor(bias)Serious | 0.5429 | 0.2088 | 2.5998 | 0.0093 | 0.1336 | 0.9523 | ** |
| age_c | -0.0116 | 0.0097 | -1.1957 | 0.2318 | -0.0307 | 0.0074 | |
| factor(vax)VV | 0.0612 | 0.2340 | 0.2614 | 0.7938 | -0.3975 | 0.5199 | |
| factor(vax)VV/mRNA | 0.1276 | 0.2384 | 0.5353 | 0.5925 | -0.3396 | 0.5948 | |
| **Signif. codes: 0 '***' 0.001 '**' 0.01 '*' 0.05 '.' 0.1 ' ' 1** | | | | | | | |

**Figure S14. Meta-regression outputs. Meta-analysis on the overall risk of Omicron infection after one booster dose: a) Random effect (RE) model.** The overall estimate is expressed as Log odds ratio and 95%CI ($\hat{\mu}$= -1.2157 (95%CI: -1.4854 to -0.9460)), gives OR= 0.296 (95%CI: 0.226 to 0.388), by exponentiation. The amount of heterogeneity in the baseline RE model is estimated to be tau$^2$ = 0.4686. The Multi Level Meta-analysis (MLM) is run in order to explore 'severity' of Covid-19 as a potential source of heterogeneity. The effect sizes are clustered by clinical 'severity' (third level) , while the variable 'author' defines the second level of the three-level model. The same RE is assigned to effect sizes within the same group, for both grouping variables, whereas different RE are assigned to effect sizes having different grouping variable. The multilevel meta-analysis (MLM) aims to assess how the variance is distributed within and between levels. The MLM output sigma2.1 = 0.346 represents the variance 'within studies', whilst sigma2.2= 0.255 the estimated value for the 'between-study' variance. The data set includes 28 effect sizes pertaining to 3 clusters ('ANY', 'SYMPT' and 'HOSP'). The test for heterogeneity shows signicant variation between all effect sizes in the data set, since the p-value is smaller than 0.001. The overall effect size can be derived from the estimate= -1.29 (95% CI:-1.95 to -0.64, p< 0.001) corresponding to OR=0.27 (95% CI: 0.14 to 0.53), hence, similar to the estimate exhibited by the RE model except shrunk CI. Both within and between group variance are significant, since the fit of the full models are significantly better than the fit of the reduced models (p<0 .0001 and p=0.049, respectively). The distribution of variance over the three levels of the model produces a within study variance $\sigma^2$=0.346 and a between group variance $\sigma^2$=0.255 (distributed at the third level of the model). Both within-study variance (level2) and between-study variance (level 3) are significant at the log-likelihood-ratio tests.Overall, 52.6% of the overall variance can be attributed to the first level, 57.3% to the second level and 42.2% to the third level ('between-study' variance). **b) Mixed effect meta-regression output (one moderator).** By including the variable 'severity' as a predictor/moderator in the meta-regression model (with intercept), 'Any positive rt-PCR' (intercept) is considered as the reference level and the estimated (average) log risk ratio is $\hat{\beta}_0 = -0.8551$. The coefficients of the remaining levels indicate how much the estimated (average) log risk ratios diverge from that value. The coefficient representing 'symptomatic' level is not significant in this model (p >0.05). However, the test of moderators is significant ($Q_M$ (df = 2) = 10.8779, p=0.0043), hence, we can reject the null hypothesis that the three level factor as a whole is not significant. Therefore, if $\exp(\hat{\beta}_0 = -0.8551) = 0.43$ yields the estimated average OR for observations related to 'Any positive rt-PCR', the average OR for 'hospitalization' will be $\exp(\hat{\beta}_0 + \hat{\beta}_2) = \exp(-0.8551 - 1.1030) = \exp(-1.9581)$ and OR= 0.14. The risk reduction for symptomatic infection is, however, 68% lower for the booster group with respect to the unvaccinated group (OR= 0.32), while the risk reduction for hospitalization is 86% smaller than the unvaccinated group (OR=0.14; 95%CI= 0.08 to 0.24). Overall, the risk of any positive rt-PCR is 57% lower in the booster group (OR=0.43; 95%CI: 0.28 to 0.64). The test for residual heterogeneity is still significant (QE (df = 25) = 3911.0446, p<0 .0001) **c) Mixed effect multilevel meta-regression output (four moderators).** As the estimated amount of residual heterogeneity in this model is equal to tau2 =0.1037, the (0.4686 - 0.1037)/ 0.4686 = 77.87% of the total amount of heterogeneity can be solved by including these moderators. The test for residual heterogeneity is still significant (QE(df = 20) = 306.8591, p< 0.0001). The coefficients 'age_c' (values centered aroun the mean of 44.4 years) and 'vax' (indicating the primary course regimen) are not significant at p<0.05. The risk estimate is on average 72% greater for the booster group in studies with serious risk of bias (OR= exp(0.5429)=1.72; 95%CI=1.14 to 2.59), whereas it is 51% lower in studies with moderate risk of bias (OR= exp(-0.7112)=0.49; 95%CI=0.31 to 0.77) compared to studies with low risk of bias (reference). The test of Moderators in the model with intercept is significant [QM(df = 8) = 350.4571, p< 0.0001].

#### SARS-COV2 INFECTION RISK AFTER VACCINATION BY SEVERITY, OR[95%CI]

| | Meta-regression OR[95%CI] | | | Multiple meta-regression OR[95%CI] | | |
|---|---|---|---|---|---|---|
| Severity | OR | LCI | UCI | OR | LCI | UCI |
| **Sars-Cov2 infection risk after primary vaccination** | | | | | | |
| Any positive rt-PCR | 1.074 | 0.828 | 1.393 | 1.302 | 0.894 | 1.898 |
| Hospitalization | 0.413 | 0.309 | 0.551 | 0.497 | 0.343 | 0.720 |
| Symptomatic | 0.681 | 0.559 | 0.828 | 0.755 | 0.576 | 0.991 |
| **Sars-Cov2 infection risk after one booster** | | | | | | |
| Any positive rt-PCR | 0.425 | 0.282 | 0.641 | 0.333 | 0.216 | 0.513 |
| Hospitalization | 0.141 | 0.084 | 0.238 | 0.121 | 0.078 | 0.186 |
| Symptomatic | 0.322 | 0.230 | 0.452 | 0.306 | 0.233 | 0.402 |

*Note:*

[1] LCI: lowerbounds of the corresponding 95% confidence intervals;

[2] UCI: upper bounds of the corresponding 95% confidence intervals.

**Table S4. ODDS RATIOs.** Estimates of meta-regression model and multiple meta-regression model by clinical severity of Omicron infection. Primary course vaccination and booster ORs [95%CI].

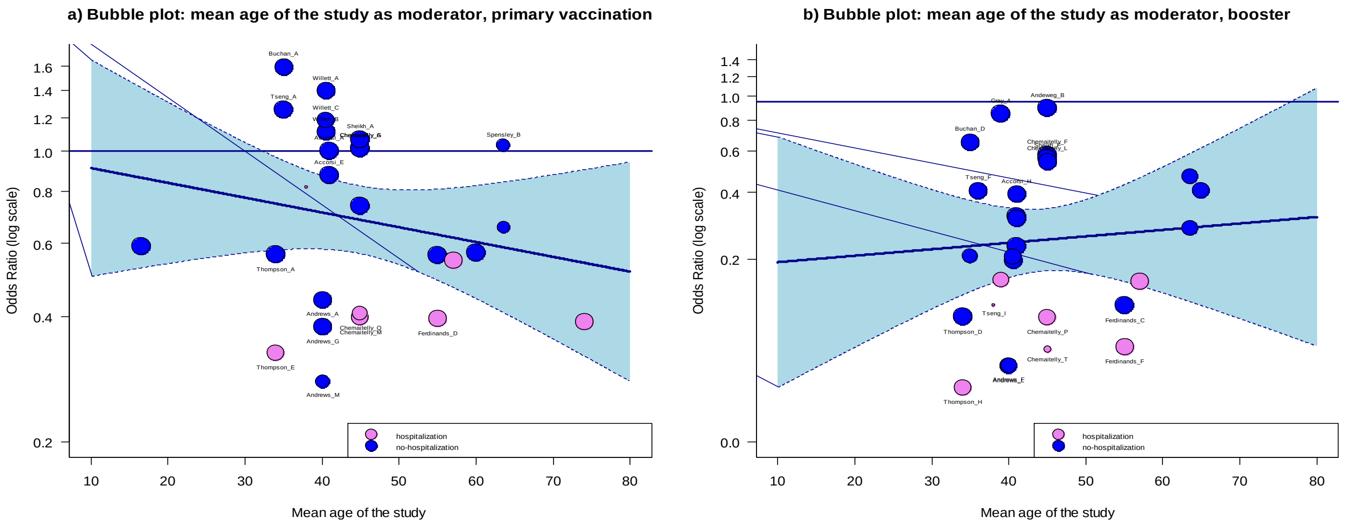

**Figure S15. Bubble plots.** The scatter plots show the observed ORs of each individual observation plotted against a quantitative predictor (mean age of the study sample). In particular, the OR estimatites for hospitalization (lighter colour) are distinguished from symptomatic and any positive rt-PCR estimates (darker colour). The size of the points is drawn proportional to the weight that the studies received in the analysis (larger points for the observations that received more weight). The regression line from the model (with corresponding confidence intervals) is added at the centre of the plot. **(a) Overall Sars-Cov2 infection risk after primary vaccination.** The mixed effect meta-regression with one moderator (and intercept) does not produce significant results for 'age' coefficient (Log OR= -0.0084; 95%CI: -0.0246 to 0.0082). The overall estimate shows, however, a downward sloping. The bubbles indicating the hospitalization ORs are quite scattered and concentrated at the bottom right of the graph, whilst the bubbles indicating the risk of any clinical Omicron infection, except need for hospitalization, appear more sparse. Although the overall risk appears to decrease with age, the ORs estimates for hospitalization slightly increase, especially around the mean values ranging from 45 to 60 years. However, the test of Moderators is not significant ($Q_M$(df = 1) = 0.1288, p = 0.7197) **(b) Overall Sars-Cov2 infection risk after one booster dose .** The mixed effect meta-regression with one moderator (intercept) does not produce significant result for 'age' coefficient (Log OR= 0.0057; 95%CI: -0.0254 to 0.0368). The overall estimate shows an upward sloping. The bubbles indicating the hospitalization ORs are less scattered and centered at the bottom of the regression line, whilst the bubbles indicating the risk of any clinical Omicron infection appear distributed above the regression line. Although the overall risk appear to increase with age, the risk estimates for hospitalization remain stable.

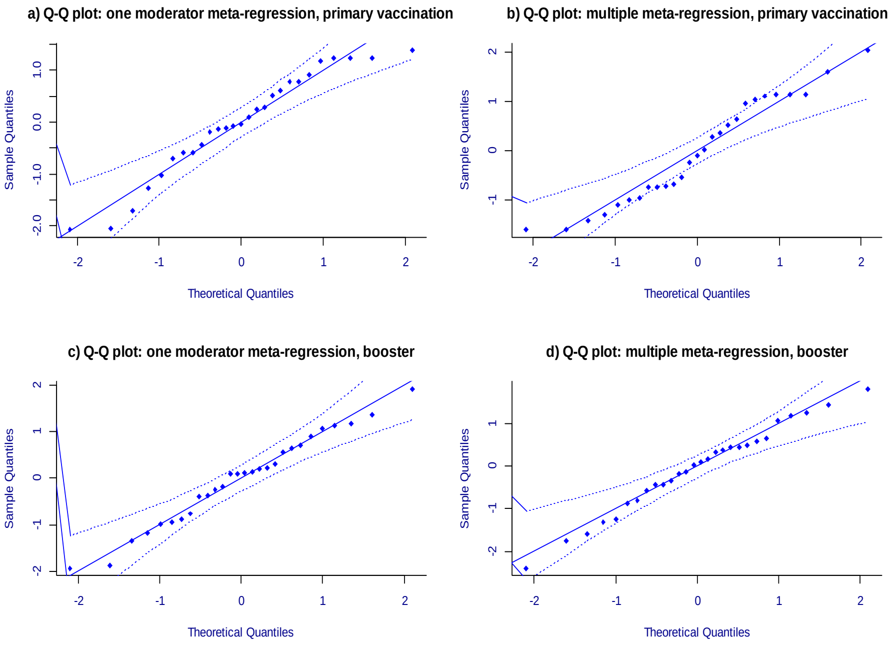

**Figure S16. Q-Q plots: mixed effect meta-regression models (REML method) on effectiveness of Sars-Cov2 primary course vaccination (a, b) and booster (c, d) against Omicron.** The normal quantile-quantile (Q-Q) plots are used in meta-analyses to check some assumptions of the data and to provide a graphical assessment of "goodness of fit". The theoretical quantiles (also known as the standard normal variate, i.e., a normal distribution with mean=0 and standard deviation=1)are plotted on the x-axis while the ordered values for the random variable(s) with the unknown distribution are plotted on the y-axis. The envelope is created basing on the quantiles of sets of pseudo residuals simulated from the given model. The number of simulated sets is 1000, by default. The simulated bounds are smoothed with Friedman's Super-Smoother. (Wang & Bushman, 1998; Cook & Weisberg, 1982) Theoretically, the points in the plot should fall on a diagonal line with slope of 1, going through the (0,0) point. Deviations indicate that the (residual) heterogeneity in the true effects is non-normally distributed, and that the residual unobserved effect is not adequately controlled by any moderators already included in the model. In **a)** and **c)** clinical 'severity' is included as moderator; while three additional predictors are added to the models (risk of bias, vaccine regimen and mean age of the study sample) in **b)** and **d).** In the mixed-effect meta-regression with one moderator (**a** and **b**), the points fall along the line in the middle of the graph, but they curve off in the extremities, suggesting more extreme values than would be expected from a normally distributed data. Moreover, although extreme values benefit from the multiple meta-regression model **(b),** some central points fall outside the envelope. The multiple meta-regression on the booster dose effectiveness shows that the points are evenly aligned with the standard normal variate **(d).**

##### *3.2 Secondary outcomes: waning effectiveness of primary course vaccination against the risk of symptomatic Covid-19 and hospitalization*

| Characteristics | Total (N=40) | Severity Symptomatic (N=29) | Severity Hospitalized (N=11) |
|---|---|---|---|
| **Study design** | | | |
| case-negative control | 40 (100 %) | 29 (100 %) | 11 (100 %) |
| **Country** | | | |
| Qatar | 12 (30 %) | 8 (28 %) | 4 (36 %) |
| UK | 20 (50 %) | 16 (55 %) | 4 (36 %) |
| US | 8 (20 %) | 5 (17 %) | 3 (27 %) |
| **Sample age (years)** | | | |
| Mean (SD) | 43.3 (7.62) | 41.4 (6.24) | 48.3 (8.95) |
| Median [Min, Max] | 45.0 [16.5, 57.0] | 41.0 [16.5, 55.0] | 45.0 [34.0, 57.0] |
| **Elderly** | | | |
| <65 years | 40 (100 %) | 29 (100 %) | 11 (100 %) |
| >65 years | 0 (0 %) | 0 (0 %) | 0 (0 %) |
| **Study follow-up period (days)** | | | |
| Mean (SD) | 209 (53.6) | 195 (47.7) | 244 (53.9) |
| Median [Min, Max] | 213 [70.0, 330] | 175 [70.0, 330] | 223 [155, 308] |
| **Induction time (days)** | | | |
| Mean (SD) | 18.1 (8.16) | 18.4 (7.28) | 17.3 (10.5) |
| Median [Min, Max] | 14.0 [7.00, 30.0] | 14.0 [14.0, 30.0] | 14.0 [7.00, 30.0] |
| **Time interval since last dose** | | | |
| 3 months | 9 (22 %) | 7 (24 %) | 2 (18 %) |
| 3 to 6 months | 7 (18 %) | 6 (21 %) | 1 (9 %) |
| 6 months | 12 (30 %) | 8 (28 %) | 4 (36 %) |
| More than 6 months | 12 (30 %) | 8 (28 %) | 4 (36 %) |
| **Primary vaccination course** | | | |
| BNT162b2 | 15 (38 %) | 10 (34 %) | 5 (45 %) |
| BNT162b2/mRNA-1273 | 7 (18 %) | 3 (10 %) | 4 (36 %) |
| ChAdOx1-S | 4 (10 %) | 4 (14 %) | 0 (0 %) |
| ChAdOx1-S/BNT162b2/mRNA-1273 | 4 (10 %) | 4 (14 %) | 0 (0 %) |
| mRNA-1273 | 10 (25 %) | 8 (28 %) | 2 (18 %) |
| **Primary vaccination: vaccine technology** | | | |
| mRNA | 32 (80 %) | 21 (72 %) | 11 (100 %) |
| VV | 3 (8 %) | 3 (10 %) | 0 (0 %) |
| VV/mRNA | 5 (12 %) | 5 (17 %) | 0 (0 %) |
| **Risk of bias (Robins)** | | | |
| Low | 4 (10 %) | 3 (10 %) | 1 (9 %) |
| Moderate | 16 (40 %) | 14 (48 %) | 2 (18 %) |
| Serious | 20 (50 %) | 12 (41 %) | 8 (73 %) |

**Table S5.** Characteristics of observations included in the quantitative synthesis of the secondary outcome

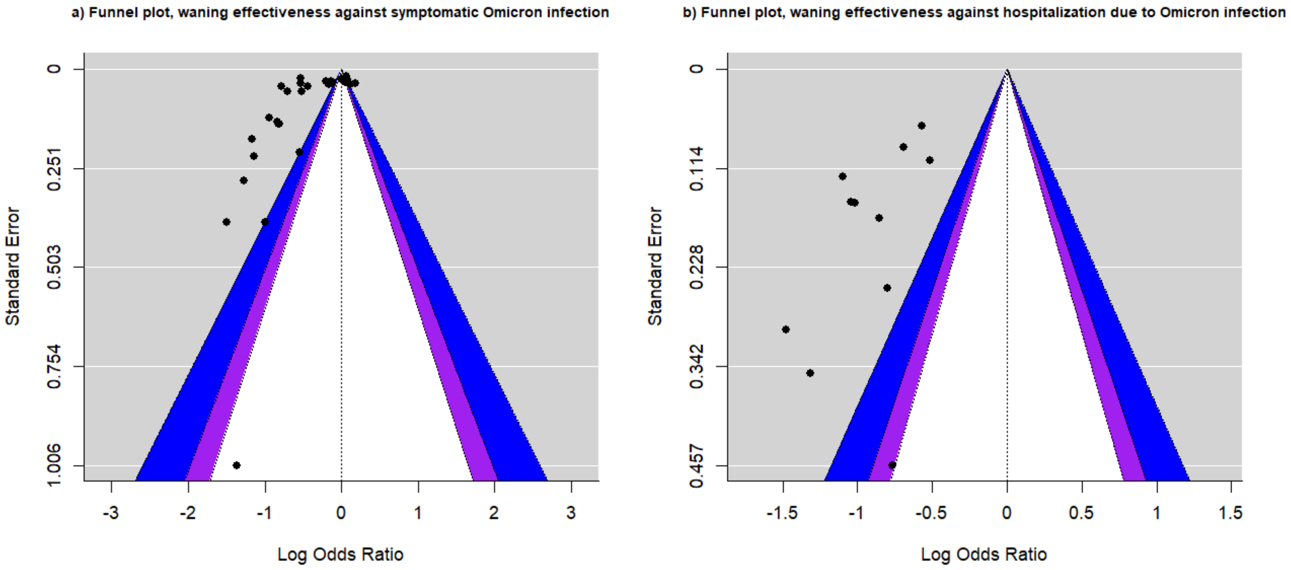

**Figure S17. Contour-Enhanced Funnel plot, effectiveness of primary course vaccination against Sars-Cov2. Symptomatic (a) and hospitalization (b) risk after primary course vaccination.** Funnel plot is used for detecting publication bias. The funnel above is centered at 0 (i.e., at the value under the null hypothesis of no effect). The three levels of statistical significance of the studies are indicated by the shaded regions. In particular, the white region in the middle corresponds to p-values greater than 0.10, the dark blue-shaded region corresponds to p-values between 0.10 and 0.05; the medium purple-shaded region corresponds to p-values between 0.05 and 0.01. Finally, the region outside of the funnel corresponds to p-values below 0.01. The plot displays the studies' results (x-axis) and their precision (y-axis). The results are expressed as log odds ratios (ORs) and the precision is represented by the standard error of the estimates. Each dot represents a single study. **a)** The larger studies tend to cluster around the top left of the plot, whilst only one smaller study is located at the bottom of the plot. The regression test indicated funnel plot asymmetry (p<0.0001) but not the rank correlation test (p=0.3051). **b)** The studies are located on the left side of the indifference line and they are quite scattered. The regression test indicated funnel plot asymmetry (p=0.0272) but not the rank correlation test (p=0.5423).

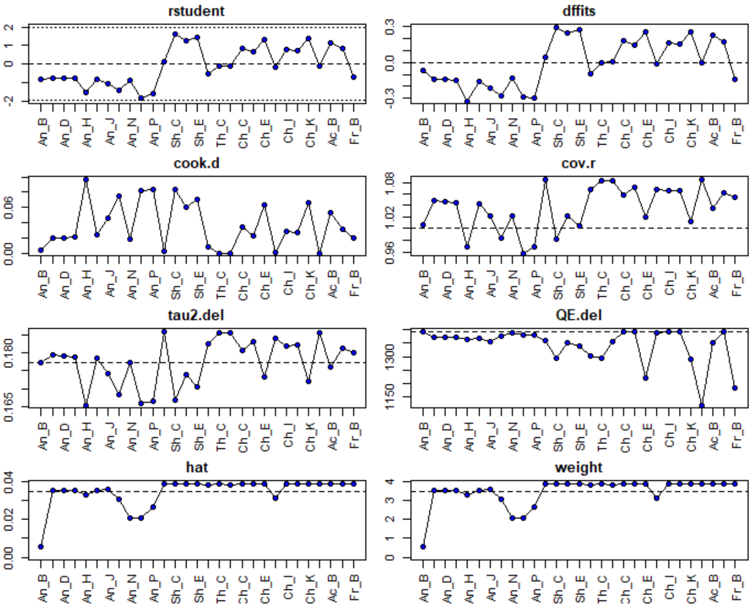

**Figure S18. Influence analysis. Waning effectiveness of primary vaccination against symptomatic Omicron infection.** According to the Q-test, the true outcomes appear heterogeneous (Q(28)=1394.3682, p<0.0001, τ2=0.1773, I2=99.0075%). A 95% prediction interval for the true outcomes is given by −1.3202 to 0.3619. Hence, although the average outcome is estimated to be negative, in some studies the true outcome may in fact be positive. The studentized residuals reveal that none of the studies has a value larger than ±3.1340 and hence there is no indication of outliers in this model. Basing on the Cook's distances, none of the studies could be considered to be influential.

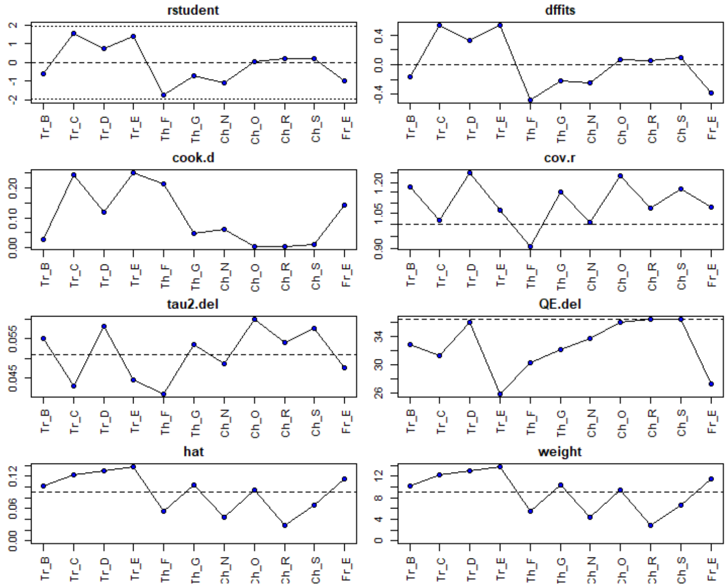

**Figure S19. Influence analysis. Waning Sars-Cov2 primary vaccination effectiveness against hospitalization due to Omicron VOC**. According to the Q-test, the true outcomes appear to be heterogeneous (Q(10)=36.3790, p<0.0001, τ^2=0.0510, I2=73.2741%). A 95% prediction interval for the true outcomes is given by −1.3381 to −0.3887. Therefore, although there may be some heterogeneity, the true outcomes of the studies are generally in the same direction as the estimated average outcome. An examination of the studentized residuals reveals that none of the studies has a value larger than ±2.8376, hence, there is no indication of outliers in the context of the model. According to the Cook's distances, none of the studies can be considered overly influential.

##### *3.2.1 Waning effectiveness: meta-regression*

**Waning vaccine effectiveness: Symptomatic Omicron infection**

```
a) Random-Effects Model (k = 29; tau^2 estimator: REML)
logLik    deviance       AIC       BIC      AICc
-18.3994   36.7987     40.7987   43.4631   41.2787
tau^2 (estimated amount of total heterogeneity): 0.1773 (SE = 0.0523)
tau (square root of estimated tau^2 value):      0.4210
I^2 (total heterogeneity / total variability):   99.01%
H^2 (total variability / sampling variability):  100.76
Test for Heterogeneity:
Q(df = 28) = 1394.3682, p-val < .0001
Model Results:
estimate      se      zval     pval     ci.lb    ci.ub
-0.4792     0.0830  -5.7742  <.0001  -0.6418  -0.3165  ***
```

```
b) Mixed-Effects Model (k = 29; tau^2 estimator: REML)
logLik  deviance       AIC       BIC      AICc
-16.0359   32.0719   42.0719   48.1662   45.2298
tau^2 (estimated amount of residual heterogeneity):      0.1689 (SE = 0.0531)
tau (square root of estimated tau^2 value):              0.4110
I^2 (residual heterogeneity / unaccounted variability): 98.87%
H^2 (unaccounted variability / sampling variability):   88.32
R^2 (amount of heterogeneity accounted for):            4.70%
Test for Residual Heterogeneity:
QE(df = 25) = 801.5493, p-val < .0001
Test of Moderators (coefficients 2:4):
QM(df = 3) = 4.1686, p-val = 0.2438
Model Results:
                          estimate    se     zval    pval    ci.lb    ci.ub
intrcpt                   -0.7297  0.1792  -4.0713  <.0001  -1.0810  -0.3784  ***
factor(time_since2)<6      0.1931  0.2341   0.8248  0.4095  -0.2658   0.6519
factor(time_since2)>6      0.4627  0.2329   1.9868  0.0469   0.0063   0.9192    *
factor(time_since2)3_6     0.2833  0.2529   1.1202  0.2626  -0.2124   0.7791
```

```
c) Mixed-Effects Model (k = 29; tau^2 estimator: REML)
logLik  deviance       AIC       BIC      AICc
11.7735  -23.5471    -3.5471    6.4102    20.8974
tau^2 (estimated amount of residual heterogeneity):      0.0042 (SE = 0.0025)
tau (square root of estimated tau^2 value):              0.0652
I^2 (residual heterogeneity / unaccounted variability): 65.10%
H^2 (unaccounted variability / sampling variability):   2.87
Test for Residual Heterogeneity:
QE(df = 20) = 53.5845, p-val < .0001
Test of Moderators (coefficients 1:9):
QM(df = 9) = 625.4783, p-val < .0001
Model Results:
                         estimate    se      zval     pval     ci.lb     ci.ub
factor(time_since2)<3    -0.6092  0.0620  -9.8203  <.0001  -0.7308  -0.4876  ***
factor(time_since2)<6    -0.2486  0.0616  -4.0369  <.0001  -0.3693  -0.1279  ***
factor(time_since2)>6    -0.0570  0.0459  -1.2420  0.2142  -0.1470   0.0330
factor(time_since2)3_6   -0.1730  0.0665  -2.5994  0.0093  -0.3034  -0.0426   **
factor(bias)Moderate     -0.6597  0.0781  -8.4427  <.0001  -0.8128  -0.5065  ***
factor(bias)Serious       0.1380  0.0547   2.5216  0.0117   0.0307   0.2452    *
age_c                    -0.0157  0.0037  -4.2989  <.0001  -0.0229  -0.0086  ***
factor(vax)VV            -0.0662  0.1214  -0.5449  0.5858  -0.3041   0.1718
factor(vax)VV/mRNA        0.1631  0.0453   3.5975  0.0003   0.0742   0.2520  ***
Signif. codes:  0 '***' 0.001 '**' 0.01 '*' 0.05 '.' 0.1 ' ' 1
```

Figure S20. Meta-regression outputs. Symptomatic Sars-Cov2 risk after primary course vaccination: a) Random effect model (RE) without moderator. The overall effect is expressed as Log odds ratio and 95%CI ($\hat{\mu}$= -0.4792 (95%CI: -0.6418 to -0.3165)), that by exponentiation gives OR= 0.62 with 95%CI: 0.53 to 0.73). The amount of heterogeneity in the baseline random effect model is estimated to be $tau^2$=0.1773. The multilevel meta-analysis is performed in order to explore 'time-intervals' as a potential source of heterogeneity. In the MLM the data set includes 29 effect sizes pertaining to four groups (time intervals). The same RE is assigned to effect sizes within the same group, whereas different REs are assigned to effect sizes having different grouping variable. The output sigma2 =0.168 represents the variance 'within-group' and sigma2=0.011 the estimated value for the variance 'between-group' (distributed at the third level of the model). The test for heterogeneity shows signicant variation between all effect sizes in the data set, since the p value is smaller than 0.001. The overall effect size can be derived from the estimate=-0.482 (95% CI: -0.680 to -0.285) significant at p<0.001, which gives OR=0.62 (95%CI: 0.51 to 0.75). The within study variance is significant(p<0.0001), since the fit of the full model is signifcantly better than the fit of the reduced model, however the the between groups variance is not significant. This implies that there is more sampling and within study variance than between study variance, in fact, the 93% of the total variance pertains to 'within-study' variance (level 2) while 0.98% and 6% to level 1 and level

3 respectively. **b) Mixed effect meta-regression output (one moderator).** By including 'time-interval' as a categorical predictor/moderator in the meta-regression model (with intercept), '0-3 months' is considered as the reference level. The intercept represents the estimated (average) log risk ratio for '0-3 months'. The coefficients of the remaining 'time-interval' levels indicate how much the estimated (average) log risk ratios diverge from the intercept. Excluding time-lapse '> 6 months' and the intercept, no coefficient is significantly different from 0 (p>0.05). The test of moderators is not significant ($Q_M$ (df = 3) = 4.1686, p = 0.2438), hence, the results indicate that we cannot reject the null hypothesis that the four-level factor as a whole is not significant. As '> 6 months' appears to have a significant influence on the effectiveness of the vaccine against the symptomatic Covid-19, the average log odds ratio after more than 6 months since the last vaccine dose uptake will be $\hat{\beta}_0 + \hat{\beta}_2 = -0.7297 + 0.4627 = -0.267$. Therefore, on average, the risk reduction is 23% after more than 6 months (OR=0.77; 95%CI: 0.57 to 1.03). The risk of symptomatic infection within '3 months' corresponds to an (average) OR=0.48 (95%CI: 0.34 to 0.69) with risk reduction of 52% with respect to unvaccinated, while the OR increases up to 0.59 (95%CI: 0.44 to 0.79) in six months **c) Mixed effect multiple meta-regression output (four moderators).** The estimated amount of residual heterogeneity is equal to $tau^2$ = 0.0042, suggesting that (0.1773-0.0042)/ 0.1773= 97.6% of the total amount of heterogeneity can be accounted for by including four moderators in the model. The test for residual heterogeneity is still significant (QE (df = 20) = 53.5845, p < 0.0001), indicating that other moderators not considered in the model can influence the outcome. The intercept is suppressed and the test of moderators is significant ($Q_M$ (df = 9) = 625.4783, p<0.0001). All coefficients are all significant (p<0.05) except '>6 months' level and 'VV' vaccine. The risk of symptomatic Covid-19 is 46% lower for vaccinated within 3 months (OR=0.54; 95%CI: 0.48 to 0.61), however, the risk reduction decreases to 16% in favour of vaccinated between 3 and 6 months (OR=0.84; 95%CI: 0.74 to 0.96). Overall, the risk results 22% lower for vaccinated within 6 months since the last dose (OR=0.78; 95%CI: 0.69 to 0.88). Moreover, the risk appears, on average, 15% higher for vaccinated in studies with serious risk of bias (OR=1.15; 95%CI: 1.03 to 1.28), and 48% lower in studies with moderate risk of bias (OR=0.52; 95%CI: 0.44 to 0.60) compared to studies with low risk of bias (reference). As 'age' is centred around the overall mean age of the studies (41.4 years), the OR decreases on average by 2% in studies where the mean age is one more unit away from the overall mean (OR=0.98; 95%CI: 0.98 to 0.99). Finally, the heterologous vaccination shows on average an increased risk of 18% (OR=1.18; 95%CI: 1.08 to 1.29) with respect to the mRNA vaccination regimen (reference).

**Waning vaccine effectiveness: Hospitalization due to Omicron infection**

**a) Random-Effects Model (k = 11; tau^2 estimator: REML)**

```
logLik  deviance       AIC       BIC      AICc

 -2.1205    4.2410    8.2410    8.8461    9.9552

tau^2 (estimated amount of total heterogeneity): 0.0510 (SE = 0.0357)
tau (square root of estimated tau^2 value):      0.2259
I^2 (total heterogeneity / total variability):   73.27%
H^2 (total variability / sampling variability):  3.74
Test for Heterogeneity:
Q(df = 10) = 36.3790, p-val < .0001
Model Results:
estimate      se      zval     pval     ci.lb     ci.ub
 -0.8634    0.0875  -9.8706  <.0001  -1.0348  -0.6920  ***
```

**b) Mixed-Effects Model (k = 11; tau^2 estimator: REML)**

```
logLik  deviance       AIC       BIC      AICc
 -1.3540    2.7080   12.7080   12.4376   72.7080

tau^2 (estimated amount of residual heterogeneity):     0.0456 (SE = 0.0436)

tau (square root of estimated tau^2 value):             0.2136
I^2 (residual heterogeneity / unaccounted variability): 61.21%
H^2 (unaccounted variability / sampling variability):   2.58
R^2 (amount of heterogeneity accounted for):            10.56%
Test for Residual Heterogeneity:
QE(df = 7) = 18.7323, p-val = 0.0091
Test of Moderators (coefficients 2:4):
QM(df = 3) = 3.9437, p-val = 0.2676
Model Results:

                         estimate    se     zval    pval    ci.lb    ci.ub

intrcpt                  -1.0596  0.1803  -5.8785  <.0001  -1.4129  -0.7063  ***
factor(time_since2)<6     0.0968  0.2459   0.3938  0.6937  -0.3851   0.5787
factor(time_since2)>6     0.2665  0.2237   1.1913  0.2335  -0.1719   0.7049
factor(time_since2)3_6    0.5436  0.2988   1.8196  0.0688  -0.0419   1.1291    .
```

**c) Mixed-Effects Model (k = 11; tau^2 estimator: REML)**

```
  logLik  deviance       AIC       BIC      AICc
  1.0198   -2.0397   13.9603    9.0507  157.9603
tau^2 (estimated amount of residual heterogeneity):     0 (SE = 0.0281)
tau (square root of estimated tau^2 value):             0
I^2 (residual heterogeneity / unaccounted variability): 0.00%
H^2 (unaccounted variability / sampling variability):   1.00
Test for Residual Heterogeneity:
QE(df = 4) = 1.8343, p-val = 0.7662
Test of Moderators (coefficients 1:7):
QM(df = 7) = 398.3483, p-val < .0001
Model Results:
                          estimate    se     zval    pval    ci.lb    ci.ub

factor(time_since2)<3    -1.2673  0.1464  -8.6583  <.0001  -1.5541  -0.9804  ***
factor(time_since2)<6    -0.9662  0.2242  -4.3099  <.0001  -1.4056  -0.5268  ***
factor(time_since2)>6    -0.8008  0.2134  -3.7525  0.0002  -1.2190  -0.3825  ***
factor(time_since2)3_6   -0.7644  0.2374  -3.2200  0.0013  -1.2297  -0.2991   **
factor(bias)Serious       0.0255  0.1994   0.1279  0.8982  -0.3654   0.4164
age_c                     0.0255  0.0115   2.2232  0.0262   0.0030   0.0481    *
```

**Signif. codes: 0 '***' 0.001 '**' 0.01 '*' 0.05 '.' 0.1 ' ' 1**

Figure S21. Meta-regression output. Hospitalization risk for Sars-Cov2 after primary course vaccination: a) Random effect (RE) model. The overall effect is expressed as Log odds ratio and 95%CI ($\hat{\mu}$=−0.8634 (95%CI: −1.0348 to −0.6920)), that back to exponentiation gives OR= 0.422 with 95%CI: 0.347 to 0.512. The amount of heterogeneity in the baseline RE model is estimated to be $tau^2$ = 0.0510. The multilevel meta-analysis (MLM) is performed in order to explore 'time-intervals' as a potential source of heterogeneity. The data set includes 11 effect sizes pertaining to four clusters. In the MLM, the variable authorID defines the second level of a three-level model, while four 'time-lapse' groups define the third level. The same RE is assigned to effect sizes within the same group, whereas different RE are assigned to effect sizes having different grouping variable. In the MLM output sigma2 = 0.051 represents the variance within studies and sigma2=0.000 the estimated value for the variance between studies. Thus, only the second level of the model is significant at the log-likelihood-ratio test (p= 0.0075). The test for heterogeneity shows signicant variation between all effect sizes in the data set, since the p-value is smaller than .001. The overall effect size can be derived from the estimate $\hat{\mu}$=−0.863 (95% CI: -1.058 to -0.668) significant at p<0.001, which correspond to OR=0.42 (95% CI: 0.35 to 0.51). The within study variance is significant (p=0.0075), since the fit of the full model is signifcantly better than the fit of the reduced model (one moderator), however, the between study variance is zero. This implies that there is not between study variance, in fact, the 73.3% of the total variance pertains to within study variance (level 2) while the 26.7% to level 1. **b) Mixed effect meta-regression output (one moderator)**. By including the 'time-lapse' as a predictor/moderator in the meta-regression model with intercept, '0-3 months' time interval is considered as the reference level. The intercept represents the estimated (average) log risk ratio for '0-3 months' and is the only significant coefficient in this model. The test of residual heterogeneity is significant (`QE (df = 7) = 18.7323, p=0.0091,` whilst the test of moderators is not significant ($Q_M$ (df = 3) = 3.9437, p= 0.2676). Hence, the results indicate that we cannot reject the null hypothesis that the four-level factor as a whole is not significant. After removing the intercept, all coefficients become significant. The p<0.0001 indicates that we can reject the null hypothesis that the (average) log risk ratio is zero for all three levels (output omitted). The risk of hospitalization within 3 months corresponds to an (average) OR=0.35 (95%CI: 0.24 to 0.49) and a risk reduction of 65% compared to unvaccinated, whilst the average OR becomes 0.38 (95%CI: 0.28 to 0.53) within 6 months. The OR raises to 0.45 (95%CI: 0.35 to 0.59) with risk reduction of 55% after more than 6 months. **c) Mixed effect multiple meta-regression output (three moderators).** The estimated amount of residual heterogeneity is equal to tau2=0, suggesting that (0.0510 -0)/0.0510 = 100% of the total amount of heterogeneity can be solved by including three moderators in the model. The test for residual heterogeneity is no longer significant (QE(df = 4) = 1.8343, p=0.7662). The coefficients are all significant (p<0.05) except 'Serious risk-of-bias'. The estimated average log odds ratio (of hospitalization due to Sars-Cov2 in those receiving the vaccine compared to those who did not) is -1.2673 (95% CI: -1.554 to -0.980). The test of Moderators is significant [$Q_M$(df = 7) = 398.3483, p<0.0001]. The mean age of the study sample is considered as a continuous variable and is centered around its mean. The results are stored in the variable age_c. The adjusted average effect corresponds to OR=exp(-1.2673)=0.28 (95%CI= 0.21 to 0.38) within 3 months and average risk reduction of 72% in favour of vaccinated. The average OR raises to 0.38 (exp(-0.9662) (95%CI: 0.25 to 0.59) within 6 months and to 0.45 (exp(-0.8008) (95%CI: 0.30 to 0.68) after more than 6 months. The variable 'age' (centered on the mean of 48.3 years) generates a significant coefficient, indicating that the risk of hospitalization increases on average by 2.6% by increasing of one unit the study mean age (OR=exp(0.0255)=1.026; 95%CI: 1.003 to 1.049)

SARS-COV2 INFECTION RISK WITHOUT BOOSTER BY TIME INTERVAL, OR[95%CI]

| | Meta-regression OR[95%CI] | | | Multiple meta-regression OR[95%CI] | | |
|---|---|---|---|---|---|---|
| Time | estimate | ci.lb | ci.ub | estimate | ci.lb | ci.ub |
| **Symptomatic Sars-Cov2 infection risk** | | | | | | |
| < 3 months | 0.482 | 0.339 | 0.685 | 0.544 | 0.482 | 0.614 |
| < 6 months | 0.585 | 0.435 | 0.785 | 0.780 | 0.691 | 0.880 |
| > 6 months | 0.766 | 0.572 | 1.025 | 0.945 | 0.863 | 1.034 |
| 3 to 6 months | 0.640 | 0.451 | 0.908 | 0.841 | 0.738 | 0.958 |
| **Hospitalization for Sars-Cov2 infection risk** | | | | | | |
| < 3 months | 0.347 | 0.243 | 0.493 | 0.282 | 0.211 | 0.375 |
| < 6 months | 0.382 | 0.275 | 0.530 | 0.381 | 0.245 | 0.590 |
| > 6 months | 0.452 | 0.349 | 0.587 | 0.449 | 0.296 | 0.682 |
| 3 to 6 months | 0.597 | 0.374 | 0.952 | 0.466 | 0.292 | 0.741 |

*Note:*

[1] pred= predicted values:OR;

[2] ci.lb= lower bound of the corresponding 95% confidence intervals;

[3] ci.ub= lower bound of the corresponding 95% confidence intervals.

**Table S6. ODDS RATIOs.** Estimates of meta-regressions on symptomatic and hospitalization risk after primary course vaccination.

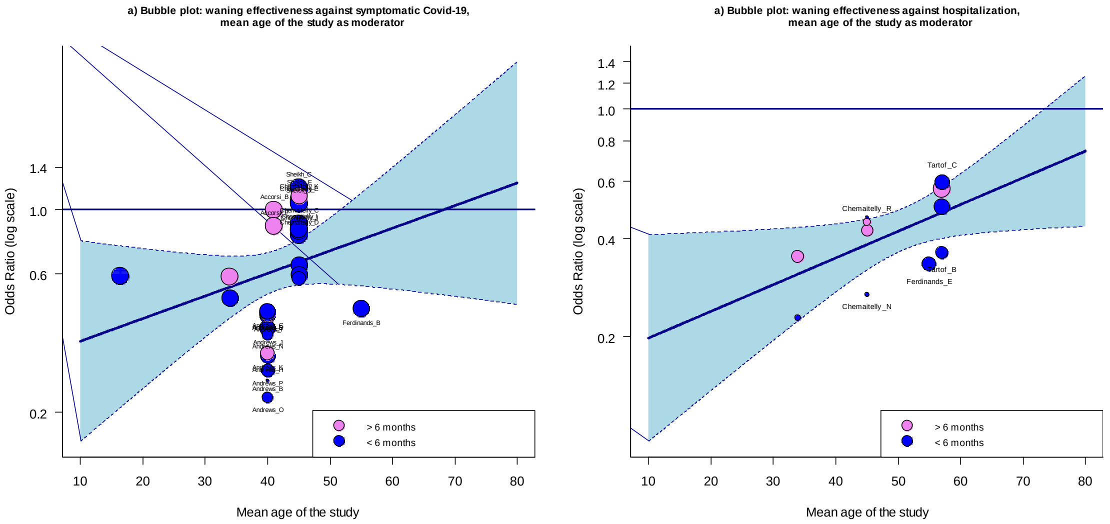

**Figure S22. Bubble plots. Waning effectiveness of Sars-Cov2 vaccination against symptomatic Covid-19 (a) and hospitalization due to Omicron infection (b).** The scatter plots show the observed ORs of each individual observation plotted against a quantitative predictor (mean age of the study sample). In particular, the OR estimatites in lighter colour represent a time interval longer than 6 months while the darker colour represents OR estimates at time interval shorter than 6 months. The size of the points is drawn proportional to the weight that the studies received in the analysis (larger points for the observations that received more weight). The regression line from the model (with corresponding confidence intervals) is added at the centre of the plot. **(a)** The mixed effect meta-regression model on symptomatic infection risk with one moderator (and intercept) does not produce significant result for 'age' coefficient (Log OR= 0.0180; 95%CI: -0.0068 to 0.0427). The overall estimate shows, however, an upward sloping. The bubbles indicating OR estimates at time interval over 6 months are scattered and concentrated at the centre of the graph mostly above the regression line, with two of them lying above the indifference line. The bubbles indicating the risk of symptomatic Omicron infection within 6 months appear more sparse and mostly concentrated below the overal estimate. Although the overall risk of symptomatic infection appears to increase with age, the ORs estimates do not show a clear pattern. The test of Moderators is not significant ($Q_M$(df = 1) = 2.0242, p=0.1548) **(b)** The mixed effect meta-regression with one moderator (and intercept) for hospitalization risk does produces significant result for 'age' coefficient (Log OR= 0.0189; 95%CI: 0.0014 to 0.0364). The test of Moderatoors is significant ($Q_M$(df = 1) = 4.4583, p=0.0347). The overall estimate shows an upward sloping. The bubbles indicating the hospitalization ORs are aligned with the regression line and the majority of them falls within the 95%CI, especially the observations relative to time intervals longer than 6 months. All the bubbles show a clear pattern, in particular, the OR values tend to be higher after 6 months and they increase with age.

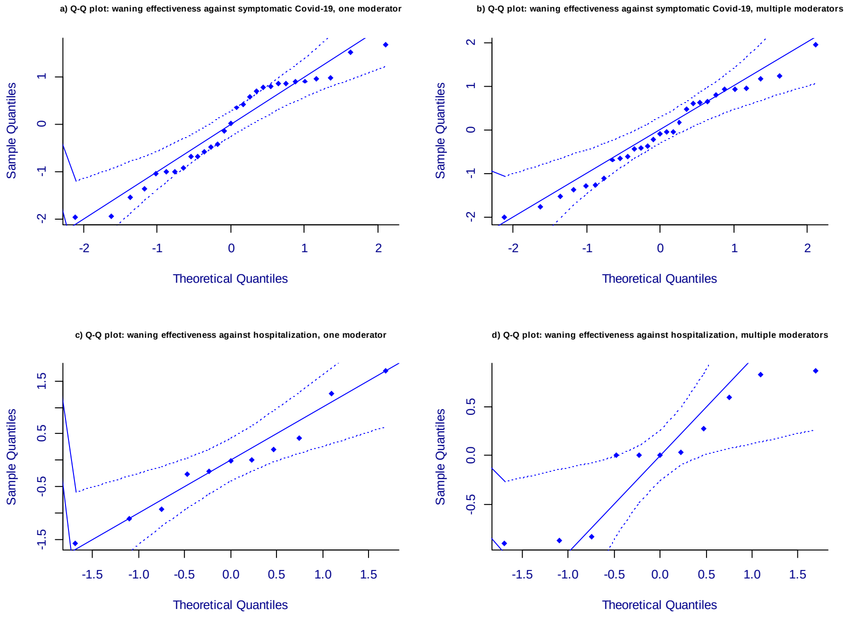

**Figure S23. Q-Q plots: mixed effect meta-regression models (REML method) on waning effectiveness of Sars-Cov2 primary course vaccination against symptomatic Omicron infection (a, b) and hospitalization due to Covid-19 (c, d).** The normal quantile-quantile (Q-Q) plots are used in meta-analyses to check some assumptions of the data and the "goodness of fit" of the meta-regression. The Q-Q plot exhibits the theoretical quantiles of a normal distribution on the x-axis against the observed quantiles of the (externally) standardized residuals on the y-axis. A reference line is added to the plot with a slope of 1, going through the (0; 0) point. The envelope is created based on the quantiles of sets of pseudo residuals simulated from the given model. The number of simulated sets is 1000, by default. The simulated bounds are smoothed with Friedman's Super-Smoother. (Wang & Bushman, 1998; Cook & Weisberg, 1982) Deviations may indicate non-normally distributed residual heterogeneity in the true effects or residual unobserved effect not adequately captured by any moderators included in the model. In **a)** and **c)** only 'time-interval' is included as categorical moderator; while three and two additional predictors are added to the models (risk of bias, vaccine regimen and mean age of the study sample) in **b)** and **d)**, respectively. The meta-regression on vaccine effectiveness against hospitalization due to Omicron is limited to mRNA vaccines. In (**a**), the points fall along the line in the middle of the graph at the extremes of the straight line, but curve off in the centre. The meta-regression benefits from three additional moderators (**b**), because the points are evenly aligned with the reference line and fall within the pseudo confidence envelope. Plots in **c** and **d** are derived from fewer observations and the point estimates appear more scattered. The multiple meta-regression does not improve the points' distribution and the envelope's bounds appear wider (**d**).
